# A Framework for Measuring Population Immunity Against Influenza Using Individual Antibody Titers

**DOI:** 10.1101/2025.07.10.25331265

**Authors:** Weijia Xiong, Xiaotong Huang, Tran Thi Nhu Thao, Nguyen Thi Le Thanh, Liping Peng, Ranawaka A. P. M. Perera, Vicky J. Fang, Bingyi Yang, Sook-San Wong, Nancy H.L. Leung, Shenna G Sullivan, Dennis K. M. Ip, Malik Peiris, Ted M. Ross, Maciej F. Boni, Benjamin J. Cowling, Tim K. Tsang

**Author notes:** **Corresponding author:** Tim K. Tsang, School of Public Health, Li Ka Shing Faculty of Medicine, The University of Hong Kong, 7 Sassoon Road, Pokfulam, Hong Kong.

## Abstract

Measuring population immunity is crucial for epidemic control and prevention of infectious diseases. While correlates of protection have been identified at the individual level for some pathogens, methods to translate these into population-level immunity metrics remain underdeveloped. We developed and validated a framework to construct population immunity estimators derived from individual serological measurements. Using influenza as a model system, we analyzed 36,150 serum samples across 4 studies covering 19 epidemics in 2009-2020, establishing four hemagglutination-inhibiting (HAI) antibody titer-based estimators: geometric mean titer, proportion of non-naïve individuals, proportion of population immune, and relative reduction in reproductive number. We found that subtype-specific relative changes of these estimators from previous seasons predicted predominant subtypes in upcoming seasons with up to 80% sensitivity and 100% specificity. In a longitudinal cohort spanning eight influenza seasons with serum collection during epidemics, we found significant negative correlations between each estimator and subsequent cumulative incidence for H1N1 viruses, but weaker correlations for H3N2 viruses. These relationships remained consistent regardless of the specific protection threshold assigned to different antibody titer levels, provided individual-level protection was at least moderate. Simulation studies revealed that lower effective reproductive numbers and higher antibody waning rates diminished the correlation strength. Our framework provides a systematic approach for evaluating population immunity estimators that could inform preparedness efforts for seasonal epidemics and potentially extend to other infectious diseases.

## INTRODUCTION

Respiratory virus infections such as influenza viruses and SARS-CoV-2 pose a substantial public health threat worldwide, leading to significant morbidity and mortality each year ^1, 2, 3 4^. Accurate measurement of population immunity is crucial for predicting the attack rate and severity of epidemics, guiding disease control and healthcare resource allocation. For example, this can prompt intensified population-wide vaccination programs or targeted vaccination in specific populations when population immunity levels are estimated to be low.

Immunity arises from two sources: natural infection and vaccination. For vaccinated individuals, near-term and medium-term immunity can be measured through vaccine effectiveness studies, assuming that the study has a sufficient follow-up duration ^5, 6^. In both vaccinated and unvaccinated individuals, serological assays from various study designs, including cohort and case-control studies, have demonstrated the importance of hemagglutination-inhibiting (HAI) antibody titers in assessing immunity against symptomatic influenza infection ^7, 8^. An HAI titer of 40 is commonly associated with approximately 50% protection against symptomatic influenza, although other aspects of immunity such as cellular and mucosal immunity likely also play a role in protection ^9, 10, 11, 12^. A key gap in seroepidemiology is the translation of individual immune responses into population-level immunity metrics, limiting the potential of serological data to be used in epidemic prediction. Developing reliable methods to quantify population immunity from HAI titers or other serological measures could improve preparedness for influenza epidemics and those of other emerging viruses.

In this study, we aim to establish a framework for constructing and evaluating population immunity metrics derived from individual HAI titers. Using data from multiple influenza seasons, we assess the validity of these estimators and examine key factors influencing their robustness. Finally, we conduct simulation studies to determine optimal sample size requirements for reliable population immunity estimates, providing practical guidance for surveillance program design. Our findings contribute to a more systematic approach for measuring population immunity and improving epidemic forecasting efforts.

## RESULTS

### Overview of this study

This study developed a framework to assess whether large data sets of individual immune measurements can be translated into and validated as population immunity metrics. Using influenza virus and HAI titers as our test case, we assessed four estimators: (1) Geometric Mean titer (GMT); (2) the proportion of non-naïve individuals; (3) the proportion of immune individuals, defined as the weighted average of protection levels across the number of individuals in different HAI titer levels; (4) the relative reduction in reproductive number defined as the proportional decrease in disease transmissibility due to population immunity (i.e., 111–11R_e_/R₀), reflecting the extent to which existing immunity reduces the transmissibility of influenza viruses. The first 3 estimators were calculated separately by age groups (children, adults and elderly), and then weighted by the age distribution of the respective study population based on local census data^13^. The fourth one was already age-weighted in the estimation process. Lower values indicate lower population immunity. We validated these estimators by assessing their ability to predict dominant influenza subtypes and their correlation with population-level incidence data. Then, we conducted SEIR model simulations to evaluate factors affecting their reliability (Figure 1). Finally, we conducted simulation studies to determine optimal sample sizes required for accurate population immunity estimation, The framework was applied to data from longitudinal cohorts in Hong Kong, Vietnam, and the USA.

**Figure 1.**
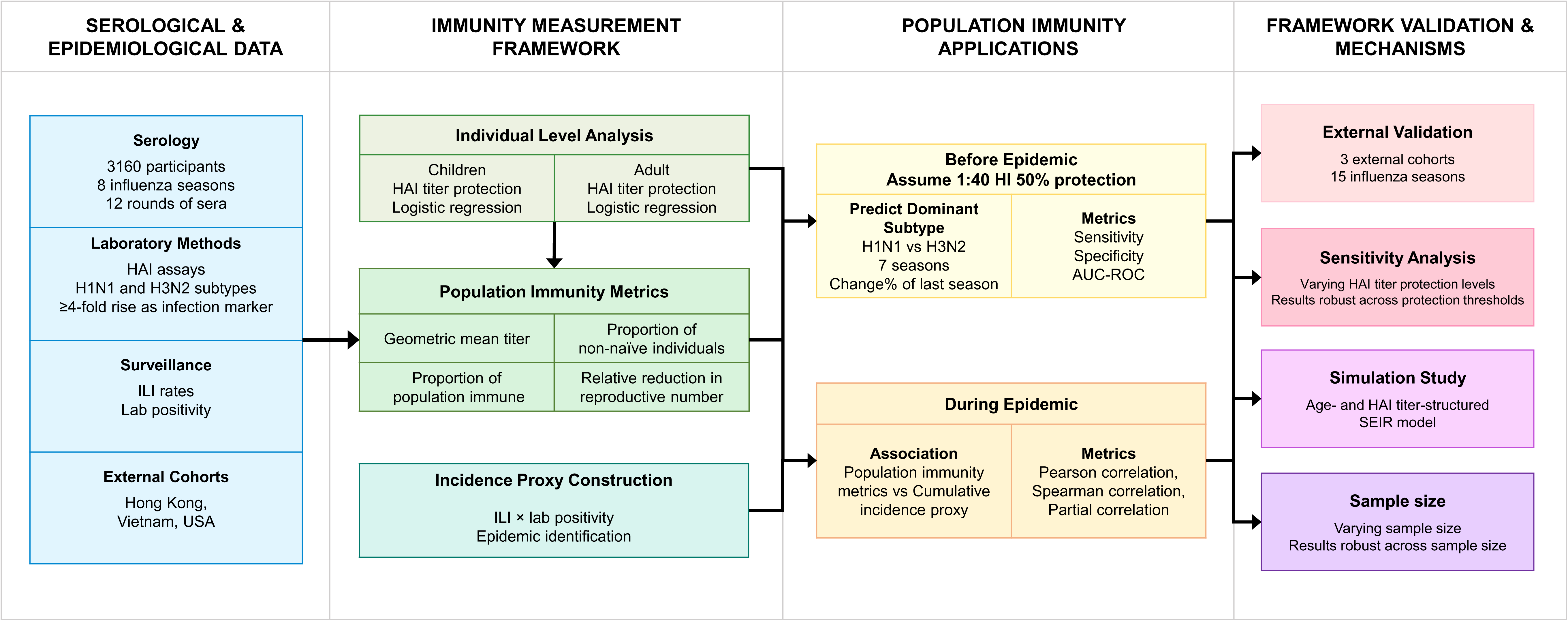
Schematic overview of population immunity measurement framework.

### Relative changes in population immunity estimators between subtypes reliably predict dominant influenza subtypes in upcoming epidemics

We computed four estimators for H1N1 and H3N2 influenza viruses using pre-epidemic titers from four studies: a Hong Kong cohort from 2009-2014^14, 15^, another Hong Kong cohort (2009-2012)^16^, a Vietnamese general population sample (2009-2012)^17^, and a general population sample from Georgia, USA (2017-2020)^18^, using the commonly adopted assumption that an HAI titer of 40 was associated with 50% protection.

We predicted the dominant subtype by tracking how population immunity changed for each subtype separately across seasons. For each subtype (H1N1 and H3N2), we calculated the season-to-season change in all four population immunity estimators. We then compared these changes between subtypes. When one subtype (e.g., H1N1) showed a greater increase in immunity than the other (H3N2), we predicted the subtype with the smaller immunity increase would dominate the following season.

For epidemics with a clear dominant subtype (>70% proportion), our four estimators showed sensitivity ranging from 60–80%, specificity from 88–100%, and AUROC from 76–94% in predicting the dominant subtype. When including all epidemics (including 2 with a dominant subtype <70% proportion), these values were 57-71%, 88-100%, and 81-90%, respectively (Figure 2A). These results support the predictive capability of relative changes (from season to season) of population immunity estimators between subtypes for predicting dominant subtypes in upcoming epidemics, though this association weakened during periods of co-circulation of H1N1 and H3N2 influenza viruses. Using season-to-season changes rather than absolute values was essential, as H3N2 influenza viruses consistently showed higher absolute population immunity levels than H1N1 influenza viruses across 15 out of 19 epidemics (Figure 2B, S1-4). Notably, our sensitivity analyses showed that varying the protective effect of an HAI titer of 40 (30% vs. 60%) had minimal impact on the predictions (Figures S5-8).

**Figure 2.**
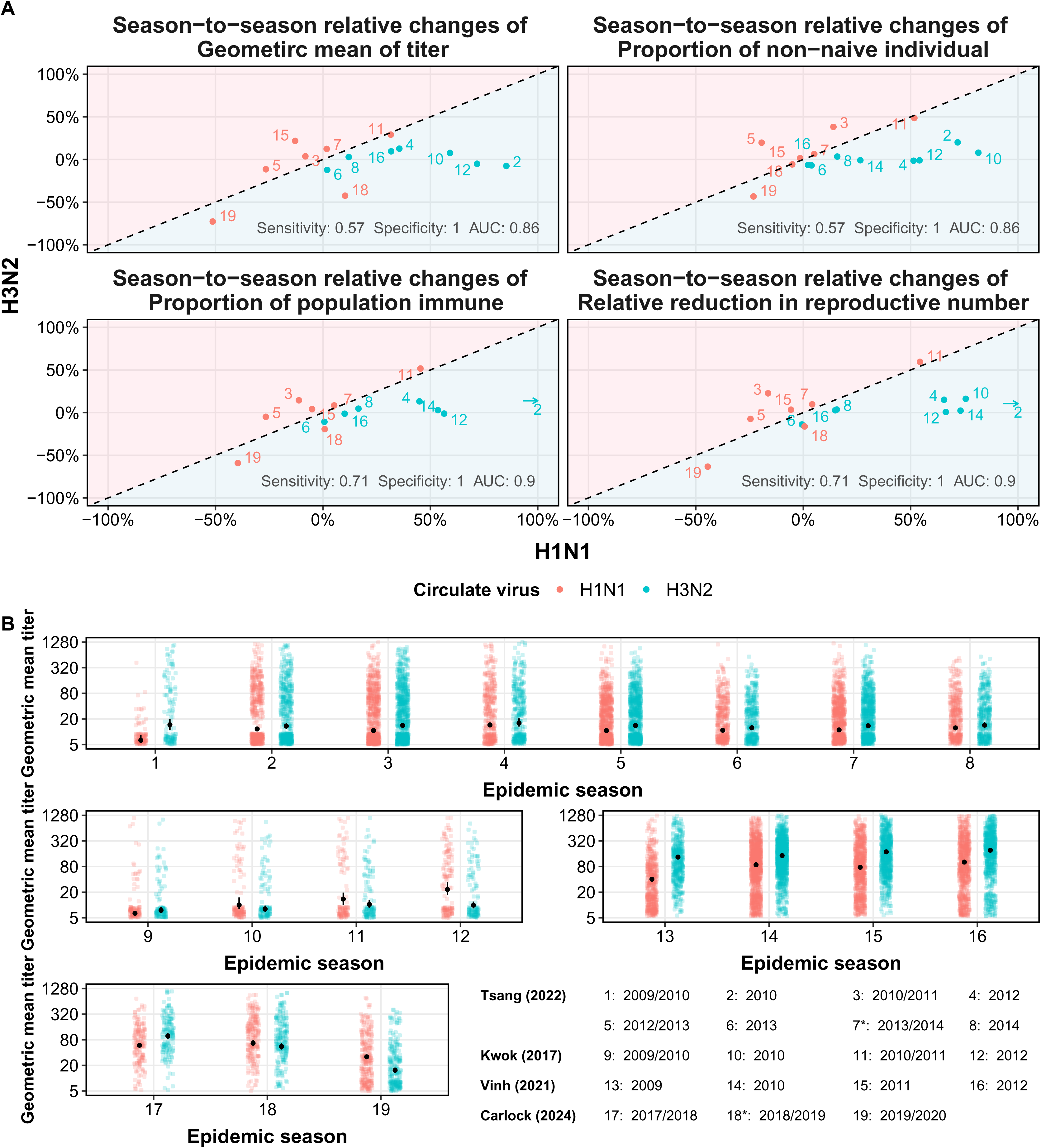
**A** The estimators of population immunity for H1N1 and H3N2 across different influenza seasons and study cohorts. The x-axis and y-axis represent the relative change from the previous season of each estimator for H1N1 and H3N2, respectively. **B** The geometric mean titer for each epidemic season from pre-epidemic titers. Each point indicates the estimated value and the solid line shows the 95% confidence interval. Each number corresponds to a specific influenza season and study, as detailed in the legend, including data from Tsang (2022), Kwok (2017), Vinh et al (2021) and Carlock (2024). Circulating virus subtypes are indicated by color coding: H1N1, H3N2. Co-circulation seasons are indicated by stars.

To assess the robustness of relative changes of population immunity estimators between subtypes with respect to strain selections, we analyzed data from two cohorts with HAI titers against multiple strains, using a diverse panel of influenza viruses beyond the vaccine strains in our primary analysis (Figures S9-10). Our findings demonstrated that all four population immunity estimators maintained similar performance regardless of the specific strain variants used, suggesting the practical flexibility and robustness of our framework for influenza subtype predictions.

### Lower pre-epidemic population immunity predicts higher infection incidence and subtype dominance

To further explore if the population immunity estimators correlate with incidence, we focused on a cohort in Hong Kong with multiple serum collections during and after epidemics^14^. In this study, a total of 3,160 individuals participated in 2008/09 ^19^ or 2009/10 ^15^ influenza seasons, with 301 participating in both years. Follow-up continued for four years, with 12 serum collection rounds (Figure 3A) covering eight major influenza epidemics (four H1N1, four H3N2). Each epidemic can be neatly bracketed by at least two rounds of serum collection. After excluding participants who were vaccinated before or during an influenza season and those with missing antibody titers, the analysis included between 567 and 2,184 eligible individuals for each epidemic studied (Table S1).

**Figure 3.**
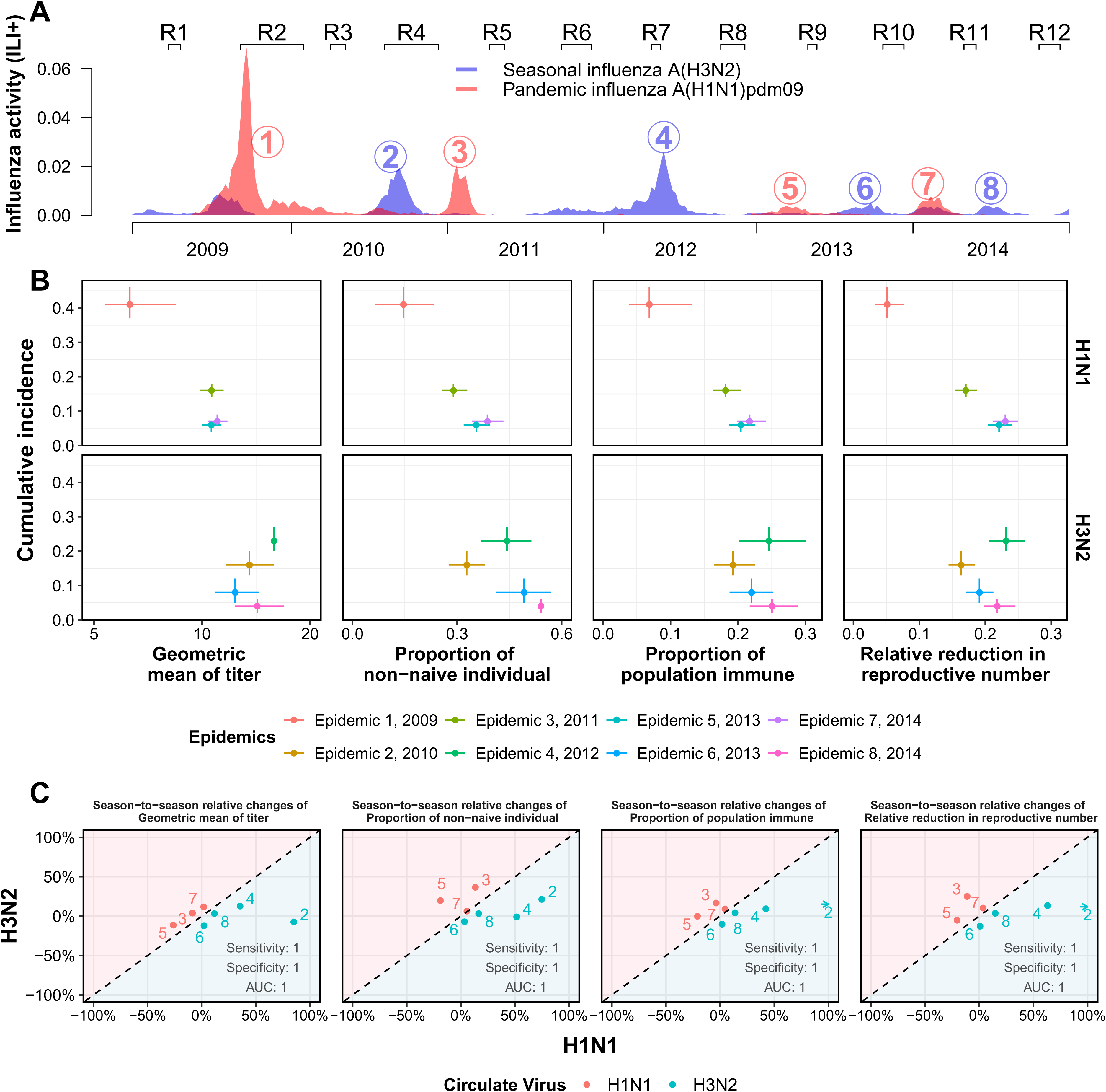
**A** Timeline of study rounds and community influenza virus activity. **B** Comparison between four estimators of population immunity and cumulative incidence of each epidemic for H1N1 and H3N2. C Comparison between four estimators of H1N1 and H3N2 before epidemics for each epidemic.

**Table 1.** Sensitivity, specificity and AUC of estimators of population immunity.

| **Estimator** | **Including epidemics with a dominate subtype only** |  |  | **Including all epidemics** |  |  |
| --- | --- | --- | --- | --- | --- | --- |
|  | **Sensitivity** | **Specificity** | **AUC** | **Sensitivity** | **Specificity** | **AUC** |
| **Geometric mean of titer** | 0.6 (0.17, 0.93) | 1 (0.6, 1) | 0.9 (0.56, 1) | 0.57 (0.2, 0.88) | 1 (0.6, 1) | 0.86 (0.59, 0.98) |
| **Proportion of non-naïve individuals** | 0.6 (0.17, 0.93) | 0.88 (0.47, 0.99) | 0.76 (0.47, 1) | 0.71 (0.3, 0.95) | 0.88 (0.47, 0.99) | 0.81 (0.58, 1) |
| **Proportion of population immune** | 0.8 (0.3, 0.99) | 1 (0.6, 1) | 0.94 (0.7, 1) | 0.71 (0.3, 0.95) | 1 (0.6, 1) | 0.9 (0.68, 1) |
| **Relative reduction in reproductive number** | 0.8 (0.3, 0.99) | 1 (0.6, 1) | 0.94 (0.7, 1) | 0.71 (0.3, 0.95) | 1 (0.6, 1) | 0.9 (0.68, 1) |

We first estimated the association between HAI titers and protection against influenza virus infection, where protection was defined as the reduction of infection risk at a specific titer level compared to HAI titers < 10 (Figure S11). For H1N1, an HAI titer of 40 provided 88% (95% CI: 86%–90%) protection for children and 60% (95% CI: 54%–65%) for adults. For H3N2, protection was 60% (95% CI: 54%–65%) for children and 51% (95% CI: 43%–58%) for adults.

Then, we calculated the four population immunity estimators based on the above estimated protections using baseline titers and excluded timeframes with <10 serum samples. A negative correlation was found between the four population immunity estimators and cumulative incidence across epidemics, except for the 2012 H3N2 influenza virus outbreak (Figure 3B).

We predicted the dominant subtype by tracking how population immunity changed for each subtype separately across seasons, using the above approach, but using estimated protection associated with HAI titers in this study. Similar to above analyses, this approach correctly classified the dominant subtype in all seven epidemics for most estimators, with only the proportion of non-naïve individuals making one incorrect prediction (Figure 3C).

### Association between population immunity estimators and infection incidence dynamics during epidemics

From the beginning to the end of epidemics, we expected these estimators to increase as population immunity accumulated throughout the course of an epidemic. Overall, we observed significant increases in estimators from pre- to post-epidemic periods. with particularly strong increases for H1N1 in epidemics 1 and 3. In contrast, epidemic 8 H3N2 exhibited a relatively constant pattern across pre- to post-epidemic periods (Figure 4),

**Figure 4.**
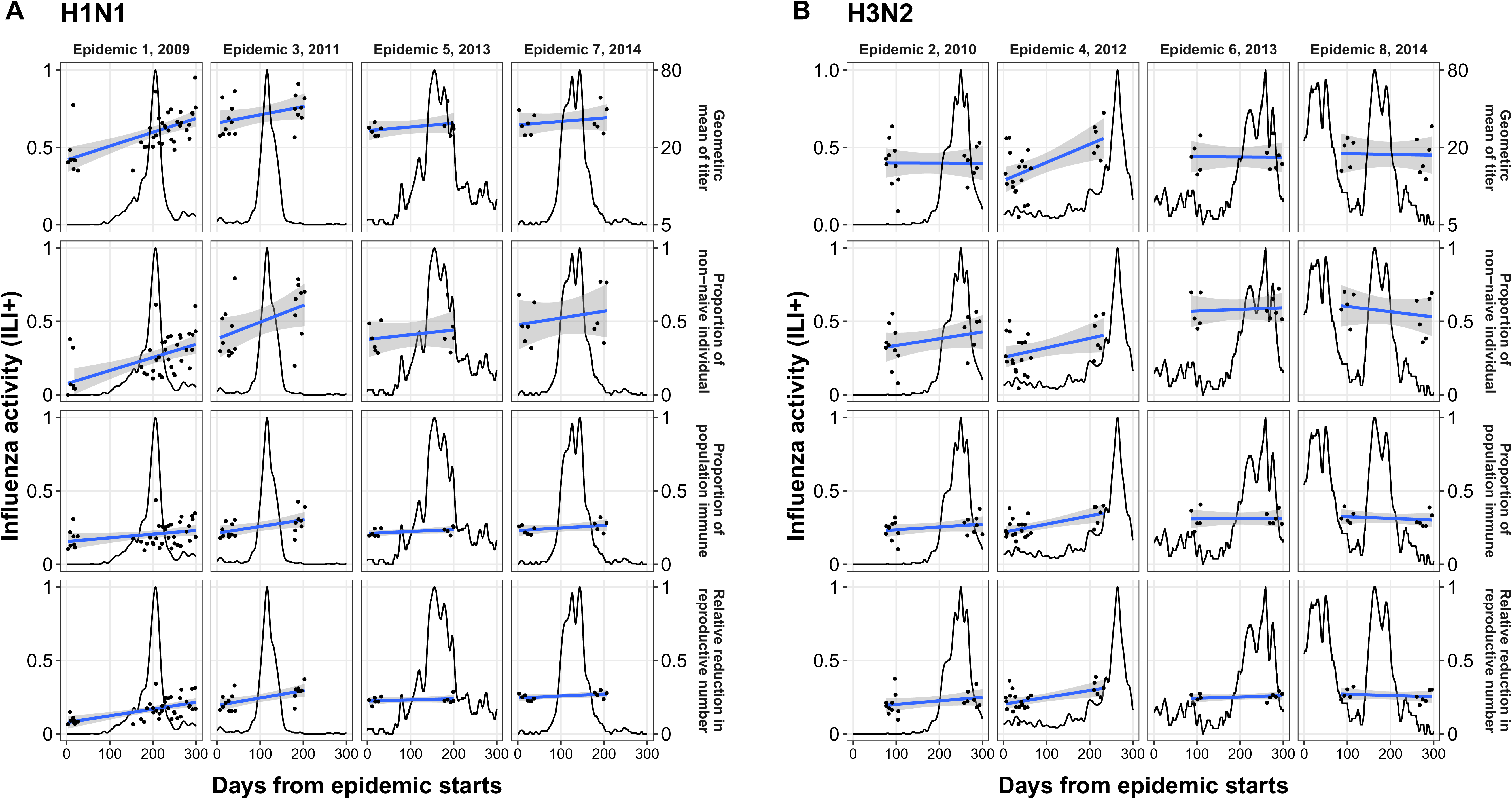
Estimators of population immunity and influenza activity proxy during each epidemic. The point represents four estimators of population immunity of each individual at that time point. The line shows the standardized influenza activity proxy in community. Panel A and B show the results of H1N1 and H3N2 from the trial cohort, respectively.

We examined correlations between the four estimators and subsequent cumulative incidence at T days following the measurement of these estimators (hereafter denoted at T-day incidence). For H1N1, three estimators showed significant negative Pearson correlations with 30-day incidence: proportion of non-naïve individuals (−0.35, 95% CI: −0.53 to −0.16), proportion of immune population (−0.37, 95% CI: −0.55 to −0.19), and relative reduction in reproductive number (−0.34, 95% CI: −0.60 to −0.24). Similar negative correlations persisted for 90-day incidence, and remained significant after adjusting for epidemic trends, except for GMT (Figure 5). These findings indicate that lower immunity estimates predicted higher near-term cumulative incidence. The correlation strength remained consistent for H1N1 over time, with stronger correlations during the 2009 H1N1 pandemic (Figure S12), which had a higher estimated R_e_ of 1.76 (95% CI: 1.48–2.04) compared to other H1N1 epidemics (Figure 5C).

**Figure 5.**
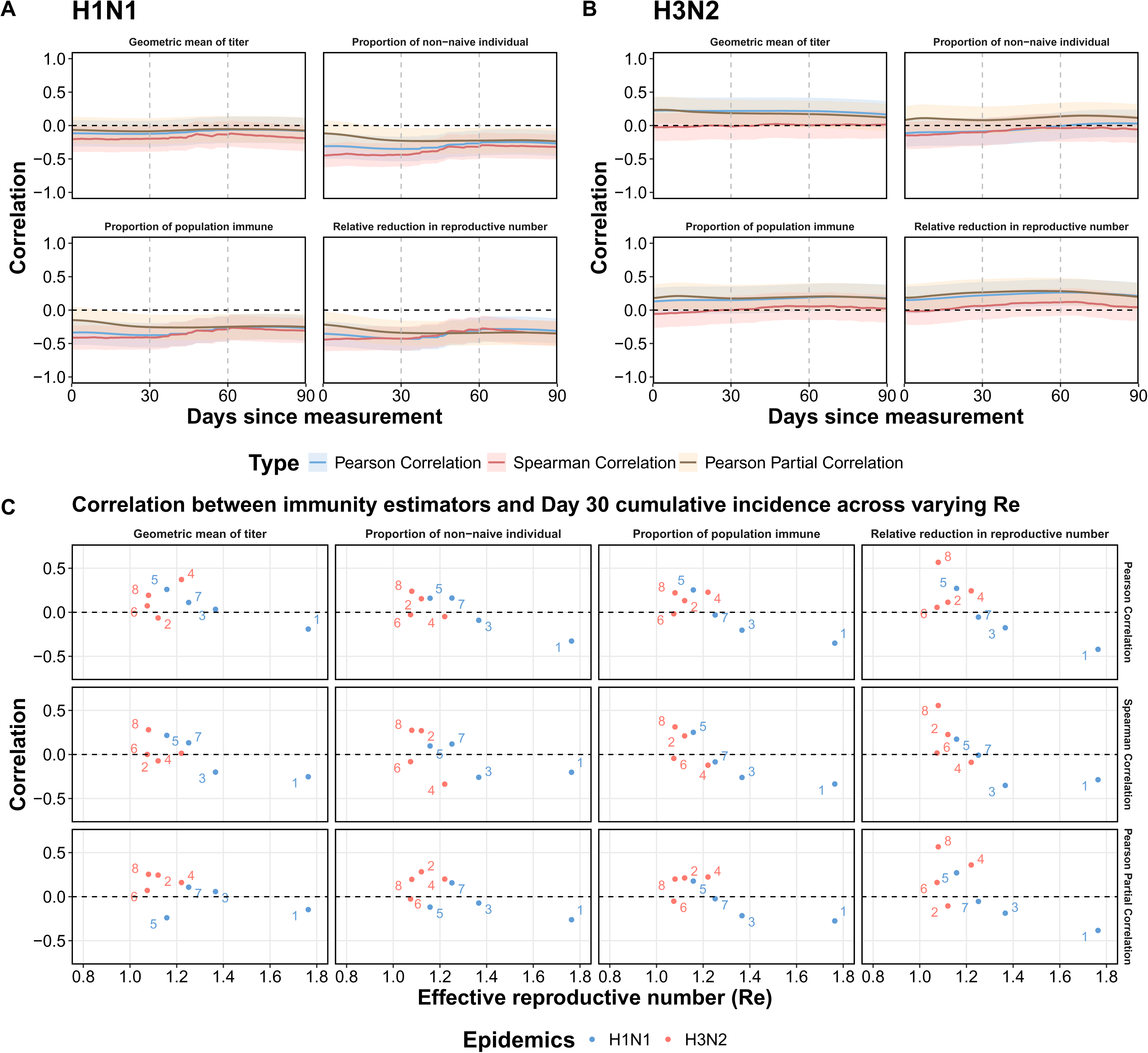
Panel A and B show the overall correlation and partial correlation of current estimators of population immunity on influenza H1N1 and H3N2 activity proxy at few days later. The blue and red lines show the Pearson correlation and Spearman correlation. The brown line shows the Partial Pearson correlation adjusted for influenza activity at previous 14 days. The shaded area represents the 95% confidence interval. Panel C illustrates the correlation and partial correlation and effective reproductive number(Re) for each epidemic.

For H3N2, we found no significant negative correlations at 30 or 90 days (Figure 5B). However, during the 2012 H3N2 epidemic, which involved a strain change from A/Perth/16/2009-like to A/Victoria/361/2011-like, we observed consistent negative Spearman correlations for proportion of non-naïve individual over time, with correlations of −0.33 (95% CI: −0.62 to −0.05) at 30 days and −0.36 (95% CI: −0.64 to −0.08) at 90 days (Figure S12). This epidemic had a higher estimated R_e_ of 1.22 (95% CI: 1.18–1.26) compared to other H3N2 epidemics in our study period (Figure 5C).

Sensitivity analyses showed that varying the protective effect of an HAI titer of 40 (30% vs. 60%) had minimal impact on correlation magnitudes (Figure S13).

### Factors affecting association between estimator and incidence of infections

We conducted simulation studies to examine these associations across various scenarios of effective reproductive number, antibody boosting, and waning rates (Figure 6). Our findings revealed that higher effective reproductive numbers were associated with weaker negative correlations between estimators and cumulative incidence. Generally, these correlations remained negative when R_e_ was 1.2 or higher.

**Figure 6.**
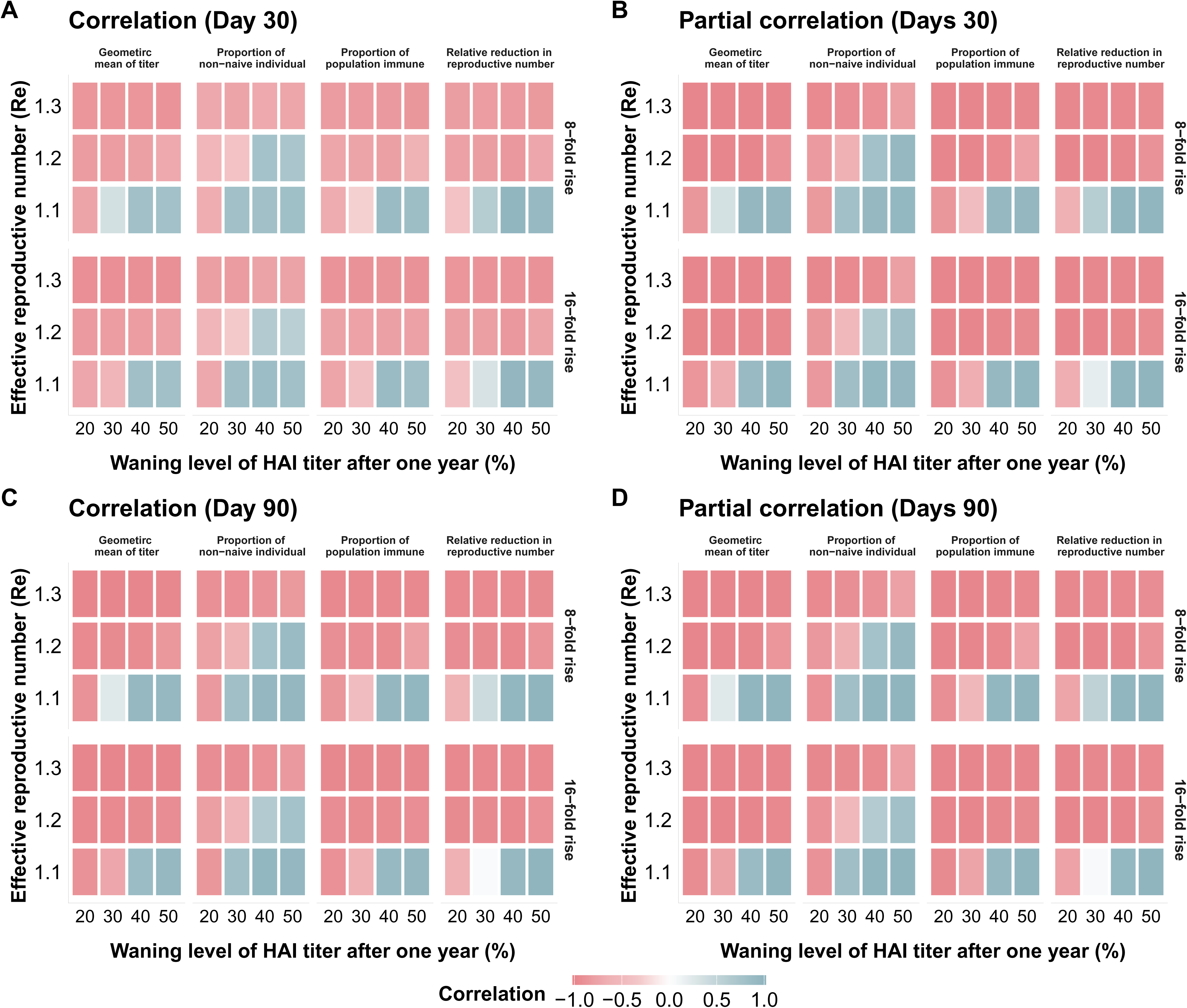
The simulation result for different effective reproductive number (Re), boosting level and waning rate of titer. This heatmap explores the correlation (Panels A and C) and partial correlation (Panels B and D) between four estimators of population immunity and cumulative influenza incidence over 30 and 90 days under various scenarios. Columns represent different waning levels of HAI titers after one year (20%-50%), and rows represent effective reproductive numbers (Re) ranging from 1.1 to 1.3. Panels A and B show results for a 30-day interval, while Panels C and D depict the 90-day interval. Separate analyses are presented for children, adults, and the overall population. Correlation values range from −1.0 to 1.0, with color intensity indicating the strength and direction of the association. Results highlight the influence of waning rates and transmissibility on the utility of these estimator as measures of population immunity.

Lower antibody waning rates strengthened the negative correlation. When HAI titers waned by 30% or more after one year and R_e_ was 1.1, the association became positive. At R_e_ values of 1.2 or higher, waning rates did not alter the correlation direction, except for the proportion of non-naïve individuals estimator.

Overall, our simulations demonstrated that higher transmissibility, higher boosting rates, and lower waning rates of HAI titers strengthened these associations. In scenarios with lower transmissibility (R_e_<1.2), combined with lower boosting rates and rapid antibody waning, the correlation between estimators and influenza incidence became insignificant or positive. Notably, the magnitude of HAI titer-associated protection had minimal impact on correlation strength (Figure S14).

### Sample size requirement on population immunity estimators

In average pre-season immunity scenarios, sample size requirements varied across our four estimators. Geometric mean titer (GMT) was most stable, requiring approximately <100 participants to maintain error below 20%. The proportion of non-naïve individuals needed 100-150 participants for similar precision, while the proportion of population immune and relative reduction in reproductive number demanded larger samples of 200-300 and 500-600 participants, respectively (Figure 7). Low pre-season immunity scenarios consistently required 20-30% larger sample sizes across all estimators compared to average immunity scenarios, with this effect most pronounced for the relative reduction metric. H3N2 generally required larger samples than H1N1. When applying a 10% error threshold, sample size requirements increased by 2 to 4-fold.

**Figure 7.**
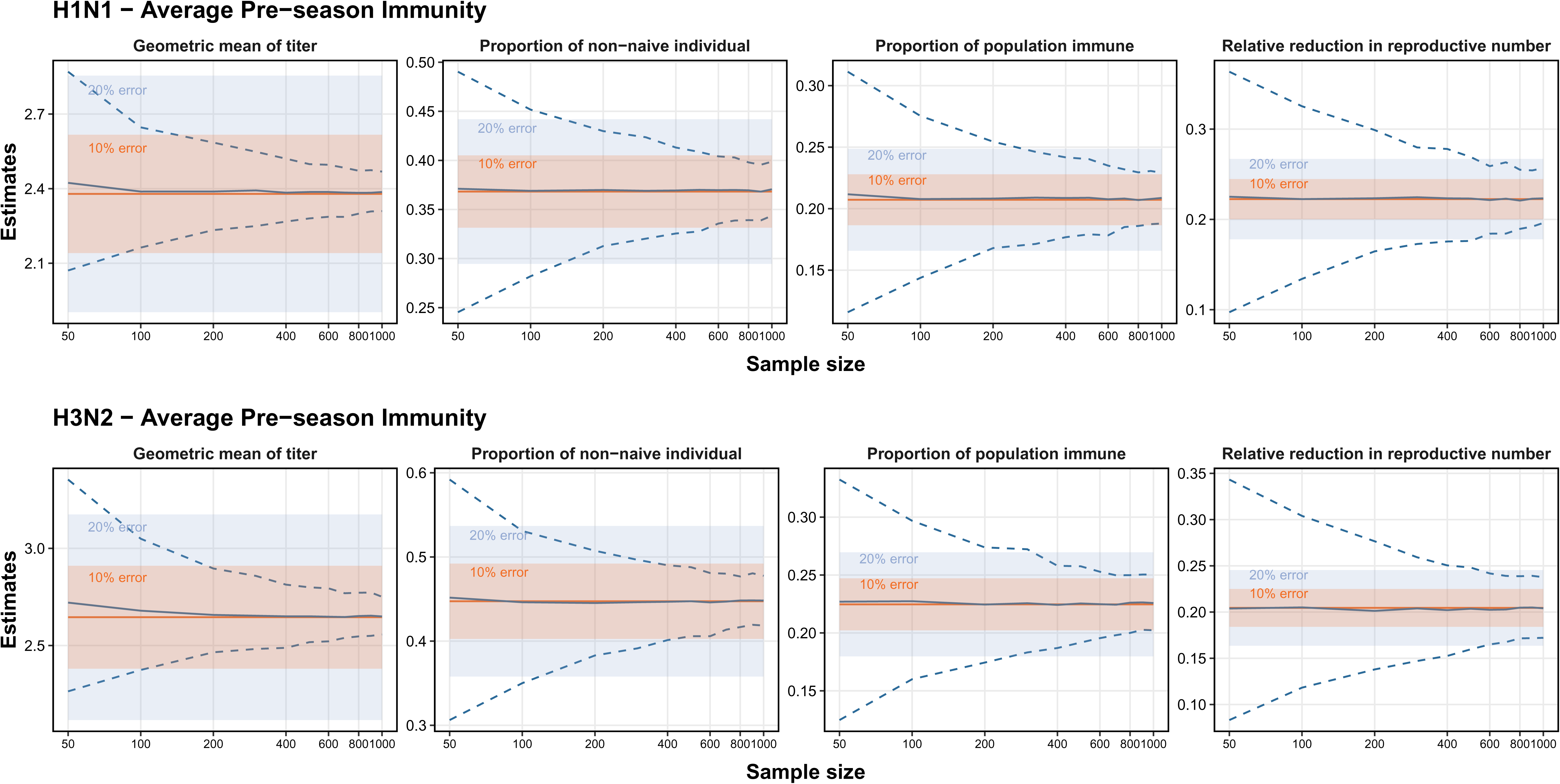
Sample size requirements for population immunity estimators with varying precision thresholds. The figure shows the minimum sample sizes required to achieve 10% and 20% error thresholds across four population immunity estimators (geometric mean titer, proportion of non-naive individuals, proportion of population immune, and relative reduction in reproductive number) for H1N1 (top panels) and H3N2 (bottom panels) influenza viruses in average pre-season immunity scenarios.

## DISCUSSION

Measuring population immunity against influenza is critical for effective disease control and prevention. In this study, we present a framework for constructing and validating population immunity derived from individual immunity measurements. Using influenza virus as an example, we evaluated whether hemagglutination inhibition (HAI) titers, the most widely used individual-level protection metric, can serve as a reliable indicator of population immunity. To do this, we developed four estimators: the geometric mean titer, the proportion of non-naïve individuals, the proportion of the population considered immune, and the relative reduction in reproductive number.

Regarding prediction of dominant influenza subtypes in upcoming epidemics, we found that the absolute values of each of the four estimators are generally higher for H3N2 than H1N1 influenza virus across most epidemics. However, higher titer for a given subtype did not necessarily correspond a higher epidemic attack rate and hence higher population immunity from natural infections for that subtype when comparing across subtypes. Possible reasons include (1) H3N2 infections may eliciting stronger antibody responses than H1N1 ^20, 21^, (2) higher evolutionary rate for H3N2 than H1N1 so that effective population-level antibody protection for H3N2 waned more quickly ^22, 23, 24, 25^, (3) mutations in H3N2 strain may affect the reliability of HI assays^26 27^. This highlights the importance of using season-to-season relative changes, rather than absolute immunity levels, when comparing subtypes to more reliable prediction of dominant strains. The relative changes approach demonstrated robust predictive power (with AUROC values of at least 81%) in determining the dominant subtype in upcoming influenza epidemics when applied to data from our cohort study with 3 additional cohorts spanning 15 influenza seasons. These results underscored strong discriminative capability of the method and its potential for predicting which subtype will drive the future epidemics. Furthermore, this methodology effectively adapted to regional circulation history, providing more contextually relevant predictions. Notably, these predictions remained consistent regardless of the specific strain variants used in HAI testing and even when the level of protection associated with HAI titers was varied, suggesting that the estimators are relatively insensitive to the magnitude of protection, provided that HAI titer is at least a moderate correlate of risk.

We evaluated the correlation between our estimators and population-level influenza incidence using serological data collected at multiple timepoints during eight influenza seasons in Hong Kong. For H1N1 influenza viruses, a significant negative correlation was observed between the estimators and subsequent cumulative influenza incidence, indicating that lower population immunity (as measured by our estimators) was associated with higher influenza incidence. In contrast, the correlation between HAI titers and influenza incidence for H3N2 influenza viruses was consistently weaker across all four estimators.

This discrepancy can be attributed primarily to more rapid antigenic drift for H3N2 compared to H1N1 ^22, 25^. This accelerated evolution creates two significant methodological challenges. First, the greater antigenic diversity of circulating H3N2 viruses means that using a single antigen in HAI assays is often insufficient for accurately assessing population protection^28^. Second, the antigens selected for these assays, frequently based on vaccine strains, are more severely affected by egg adaptations in H3N2 than in H1N1, further reducing their match to circulating viral variants ^29 30^. These factors, combined with the resulting faster waning of cross-reactive antibody titers^21, 23^, explain why our HAI-based estimators showed diminished correlation with actual H3N2 infection incidence.

Despite the overall negative correlation between the estimators and influenza incidence for H1N1 influenza viruses, considerable variability was observed among different epidemics, suggesting that multiple factors influence this relationship. Simulation studies indicated that a lower effective reproductive number (R_e_) could reduce the strength of the correlation. This finding aligned with our observation that the correlation was particularly robust and stronger during the 2009 H1N1 pandemic (epidemic 1), a notable example of a new variant/subtype entering the population. During this pandemic, very few individuals under the age of 55 had pre-existing immunity ^31^, which contributed to a higher R_e_ and a more pronounced correlation between population immunity and incidence. Additionally, the simulations highlighted that rates of antibody boosting and waning significantly impact these correlations, emphasizing the importance of transmissibility and antibody dynamics when applying these estimators as metrics of population immunity. Future research should refine the HAI-titer-based framework by incorporating these transmission and antibody dynamics parameters to enhance the precision and reliability of population immunity measurements.

HAI titer is an imperfect measure of individual immunity; for example, a titer of 40 is associated with only 50% protection, and HAI-associated protection tends to be more pronounced in children than in adults ^8, 21^. Our framework also revealed these limitations at the population level, with the correlation between estimators and actual infection incidence varying across epidemics and age groups. Alternative immunity measurements, such as HA stalk antibodies, full-length HA antibodies, NA antibodies, or T-cell responses, should be considered^9, 10, 11, 12 32^. The framework is designed to accommodate additional measures as they become available. The absence of these complementary immunity metrics may partly explain the inconsistent and moderate correlations observed in this study. Although our example focused on influenza virus and the well-established individual-level immunity measure of HAI titer, the framework can be applied to abovementioned immunity metrics with the corresponding protection data. Similarly, the framework could be adapted for other infectious diseases, such as SARS-CoV-2, using relevant immunity measures like neutralizing antibody levels ^33, 34^.

Regarding implementation of the developed framework, it requires serological sampling stratified by age groups, HAI titer testing against circulating or vaccinated strains, and calculation of the estimators. Our results suggest that using relative changes in population immunity estimators between subtypes were essential, therefore, data from one season are required to be served as baseline. To facilitate implementation of these methods, we developed an R package that allows researchers to calculate and validate population immunity metrics using their own serological datasets.

This study has several limitations. First, we relied on an influenza proxy to represent community-level incidence. Although validated against hospitalization epidemic curves, the accuracy of our results depends on the reliability of this proxy. Second, we used serological data to infer infections based on fourfold or greater rises in HAI titers, which may have led to missed infections or misclassification due to measurement error ^35^. Third, vaccination status was self-reported, which may have introduced misclassification by including individuals with HAI titer rises due to vaccination rather than infection (with the exception of Vietnam where influenza vaccine coverage at the time of sample collection was known to be lower than 1% ^36^). Additionally, the timing of serum collection did not consistently bracket each community epidemic, potentially introducing variability^37, 38^.Finally, we did not account for the cross-reactivity of antibodies, which may have influenced both HAI titers and the observed protection ^39^.

In conclusion, our study developed and validated a framework based on four population immunity estimators. Using influenza as a model, we demonstrated that these HAI titer-derived estimators correlate negatively with subsequent cumulative influenza incidence for H1N1 influenza viruses, although challenges remain for H3N2 influenza viruses due to their rapid evolution. The predictive ability of relative changes of these estimators between subtypes for identifying dominant subtypes in upcoming seasons suggests that serological surveillance can play an important role in seasonal influenza preparedness, particularly when integrated with other surveillance methods. Future work should focus on incorporating multiple immunity measures, addressing the limitations of HAI titers, and refining methodologies to account for dynamic antibody waning and viral evolution to improve public health interventions and adaptive strategies against influenza outbreaks.

## METHODS

### Data sources

We first used data from two community-based randomized trials of inactivated influenza vaccination conducted in Hong Kong between 2008 and 2010, with follow up through 2014, to develop our analytical framework. To validate this framework, we incorporated additional datasets, including another longitudinal population-based serological survey conducted in Hong Kong from 2009 to 2012; a longitudinal serological cohort from Georgia, USA from 2016 to 2022; and a cross-sectional serological survey from Vietnam from 2009 to 2012.

#### Data from two community-based randomized trials on inactivated influenza vaccination in Hong Kong between 2008-10

Data were collected from two community-based randomized controlled trials (RCTs) designed to evaluate the direct and indirect benefits of influenza vaccination. Between November 2008 and October 2009^19^, 119 households were enrolled. Between September 2009 and January 2010^15^, 796 households were enrolled. In both trials, one child aged 6–17 years per household randomly assigned to receive either the trivalent inactivated influenza vaccine (TIV) or a saline placebo. Children and household members were followed for up to 5 years post-vaccination^40^. Sera were collected from all participants immediately before vaccination (pre-vaccination), one month after vaccination (post-vaccination), and annually each autumn, with additional samples obtained at six-month intervals from 25% of participants each spring. Only one child per household received TIV or placebo at the study’s outset. Annual records were kept regarding influenza vaccinations received by other household members in the first year and by any participant from the second year onward.

Serum specimens were tested using a hemagglutination-inhibition (HAI) assay to measure antibody responses against A/California/7/2009(H1N1) and A/Perth/16/2009-like(H3N2) during the first two years, and against the same H1N1 virus and A/Victoria/361/2011-like(H3N2) in years 3–5. All specimens were tested in serial doubling dilutions starting from an initial dilution of 1:10, with antibody titers defined by the highest dilution that prevented hemagglutination. Titers below the detection limit (<10) were imputed as 5.

#### Data from a longitudinal population-based sero survey in Hong Kong between 2009-12

We utilized another dataset from a longitudinal, community-based serological study conducted in Hong Kong to calculate our four measures of population immunity ^16^. Participants were recruited through random-digit dialing of household landlines. The study comprised four rounds of serum collection, corresponding to four epidemic seasons between 2009 and 2012. Serum samples from each participant were tested using HI assays against the influenza subtypes A/California/7/2009 (H1N1) and A/Perth/16/2009 (H3N2).

#### Data from a serial cross-sectional serosurvey in Vietnam between 2009-12

Serological data from southern and central Vietnam were used to compute our four measures of population immunity for subtypes A/California/7/2009 (H1N1) and A/Wisconsin/67/2005(H3N2), A/Brisbane/10/2007(H3N2), A/Perth/16/2009(H3N2) and A/Victoria/361/2011(H3N2) during influenza seasons between 2009 and 2012. These data were collected as part of a large serum bank aimed at answering questions on general-population disease incidence, immunity, and vaccination coverage ^41 36 42 43, 44 45^. Briefly, cross-sectional general-population serum samples were collected from hospital biochemistry and hematology laboratories, on a bimonthly or 4-monthly basis, from ten hospitals in central and southern Vietnam. Data from all sites and years 2009 to 2012 were used from this study. The serological data generated through this study provide a dilution-based antibody binding measure ^41 46 47^ that correlates to but is not directly translatable to an HI titer.

#### Data from a longitudinal vaccine trial with standard or high-dose influenza vaccines in United States between 2016-22

A longitudinal serological cohort data from Georgia, USA, were used to compute our four measures of population immunity^18^. During this study, participants aged 10 to 86 years received the split-inactivated influenza vaccine (Fluzone®) across six consecutive influenza seasons, from 2016–2017 to 2021–2022. Serum samples were collected at the time of vaccination (Day 0, D0) and again 21–28 days post-vaccination (D28). For our analysis, we utilized D0 serum samples collected during three epidemic seasons from 2017–2018 to 2019–2020, including multiple influenza strain subtypes, specifically A/California/07/2009 (H1N1), A/Michigan/45/2015(H1N1), A/Brisbane/02/2018(H1N1), A/Hong Kong/4801/2014(H3N2), A/Singapore/INFIMH-16-0019/ 2016(H3N2) and A/Kansas/14/2017 (H3N2).

### Ethics

In two community-based randomized trials on inactivated influenza vaccination in Hong Kong, all participants aged 18 years and older gave written informed consent. Proxy written consent from parents or legal guardians was obtained for participants, with additional written assent from those aged 8–17 years. This study protocol was approved by the Institutional Review Board of the University of Hong Kong and by the Hong Kong Department of Health Ethics Committee (Clinical Trials Registration. NCT00792051). The ethics of longitudinal population-based sero survey in Hong Kong was approved by the Institutional Review Board of the University of Hong Kong. Serological data from Vietnam were collected as part of the 02FL post-pandemic cross-sectional seroprevalence study which was approved by the Scientific and Ethical Committee of the Hospital for Tropical Diseases in Ho Chi Minh City and the Oxford Tropical Research Ethics Committee at the University of Oxford (UK). The longitudinal vaccine trial in United States trial was approved by Western Institutional Review Board and The University of Georgia Review Board.

### Statistical Analysis

Weekly age-specific consultation rates for influenza-like illness (ILI) from Private Medical Practitioner Clinics (chp.gov.hk) were combined with the weekly proportion of sentinel respiratory specimens testing positive for influenza to create a proxy measure (ILI+) of weekly influenza virus infection incidence. This proxy, which correlates more strongly with community incidence than ILI rates or laboratory detection rates alone, was used to identify influenza epidemics during the study period ^48 49 50^. Age-group–specific cumulative incidences of infection for each H1N1 and H3N2 epidemic were estimated using Poisson regression, with cumulative influenza activity(derived from the surveillance proxy) as an offset, following a previous study ^15 40^. The model is defined as

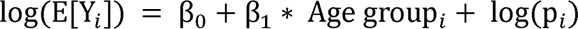

where Y_i_ is the infection number of individual i, p_i_ is the cumulative influenza activity proxy, and log(p_i_) is included as an offset in the model.

To capture the entire H1N1 wave from July 2009 to January 2010, we incorporated additional published data^19^ on H1N1 incidence among unvaccinated individuals prior to August 2009. Because enhanced surveillance during the 2009 H1N1 pandemic inflated the influenza proxy, an additional rescaling parameter^51^ was included in the regression model for data after 11 November 2009. The effective reproductive number (Re) for each epidemic was estimated from the growth rate^52^, assuming gamma-distributed serial intervals^8^ (mean 2.7 days, SD 1.0 day for H1N1; mean 2.6 days, SD 0.8 day for H3N2).

### Estimation of protection associated with HAI titers

HAI titers of 10, 20, 40 … 2560 were translated on a log2 scale to 1, 2, …, 9, with titers below 10 set to 0. Logistic regression was used to assess the association between pre-epidemic HAI titers and the risk of infection for an individual, defined as a fourfold or greater rise in consecutive sera (Appendix Section 1.1). Vaccinated individuals were excluded to isolate the effect of natural infection^53^. Analyses were conducted separately for children and adults, and for H1N1 and H3N2

### Estimators of population immunity

To estimate population immunity, we developed four key estimators: 1) geometric mean titer (GMT), 2) proportion of non-naïve individuals, 3) proportion of the population immune, and 4) relative reduction in reproductive number (Appendix).

Seroprevalence data were stratified into three age groups (0–17, 18–50, and over 50 years) and categorized across ten HAI titer levels (<10, 10, 20, 40, 80, 160, 320, 640, 1280, ≥2560). The most recent Hong Kong census data were used to define the population age distribution^54^. Bayesian inference, with Dirichlet conjugate priors for multinomial likelihoods and noninformative priors, was applied to estimate population proportion in each HAI titer group within each age stratum. GMT, proportion of non-naïve individuals, and proportion of the population immune were then calculated by adjusting the sample profile to reflect the population’s age distribution. This age-weighting process ensured that our estimators accurately represented the true demographic structure of the population rather than being biased by the age distribution of our sample.

For the relative reduction in the reproductive number, we applied the modeling framework of Cheung et al^55^. A next-generation transmission matrix was constructed using the Hong Kong social contact matrix^56^, with R₀ defined as its largest eigenvalue^57^. A modified matrix, accounting only for the susceptible population (i.e., accounting for the gradient of protection conferred by different HAI antibody titers across age groups), was used to calculate the effective reproductive number (Re), and the relative reduction in transmissibility was derived accordingly. Ninety-five percent credible intervals (CrI) for each estimator were generated at each time point and by influenza subtype through 1,000 repeated samples drawn from the joint posterior distribution for each age group (Appendix).

### Correlation between estimators of population immunity and incidence of influenza virus infection

To validate the estimators, we examined the correlation between each estimator at day t and the cumulative influenza incidence from day t to t+90, using both Pearson and Spearman correlation coefficients. Pearson partial correlation was also employed to assess the association between each estimator (St) and cumulative influenza activity (from day t to t+90), adjusting for the epidemic trend as proxied by the influenza activity 14 days before t (P_t-14_). Similar analyses were conducted for age-specific estimators (children versus adults), by epidemic, by influenza type, and by the timing of serum collection.

### Prediction of dominant subtype of upcoming season using relative changes of population immunity estimators among subtype

Since H3N2 consistently exhibited higher absolute levels than H1N1 across all epidemics for all population immunity estimators, we used subtype-specific relative changes of estimators from the previous season to current one. These changes were calculated as (X_s+1_−X_s_)/X_s_, where X_s_ and X_s+1_ were the value of estimator in season s and s+1 respectively. In principle, lower relative changes of an estimator for a given subtype should predict the subtype dominating the subsequent season. To test this, data from three additional cohorts^16, 17, 18^ were collected to evaluate predictive performance using sensitivity, specificity, and the Area Under the Receiver Operating Characteristic Curve (AUROC). A season was defined as having a dominant subtype if that subtype accounted for more than 70% of cases.

### Simulation study

An age- and HAI titer–structured SEIR model was used to investigate the relationship between the four proposed estimators and infection incidence. The total population (N = 7,150,000) was divided into three age groups (0–17, 18–50, over 50 years) and ten HAI titer level groups (<10, 10, 20, 40, 80, 160, 320, 640, 1280, ≥2560). Let *N_lk_* (*t*) denote the population in age group i with HAI titer level k at time t; these populations were further subdivided into susceptible (*S_lk_ (t)*), exposed (*E_lk_ (t)*), infectious (*I_lk_(t)*), and recovered (*R_lk_(t)*), classes. The Hong Kong contact matrix was incorporated to account for age-specific contact patterns. Detailed model descriptions and parameter specifications are provided in the Appendix and Table S2. In each simulation, the four estimators were computed, and the correlation between each estimator at day t and the cumulative infection incidence from day t to t+30 and t+90 was determined.

### Sample Size Calculation

To determine optimal sample size requirements, we conducted bootstrap simulations using serological data from the Hong Kong cohort. We selected seasons with minimum, median, and maximum GMT levels for both H1N1 and H3N2 subtypes to represent varying immunity profiles. For each scenario, we evaluated the precision of our four population immunity estimators across sample sizes ranging from 50 to 1000 participants, using 1,000 bootstrap iterations per sample size. Estimation error was quantified as the absolute percentage deviation from the true value, with a 10% and 20% error threshold as our primary criterion for adequate sample size. We performed age-stratified resampling to maintain population representativeness according to Hong Kong census data. All statistical analyses were performed using R version 4.2.2 (R Foundation for Statistical Computing, Vienna, Austria).

## DATA AVAILABILITY

All raw HAI data are publicly available from the original publications. The integrated datasets used in this study are accessible on GitHub at https://github.com/wjxiong5633/ImmuPop.

## CODE AVAILABILITY

Source code for the R scripts used in the analysis is available at https://github.com/wjxiong5633/ImmuPop. The source code of the package ImmuPop, with instructions for the package installation, is available at https://github.com/wjxiong5633/ImmuPop.

## ACKNOWLEDGMENTS

This work was supported by the Health and Medical Research Fund, Food and Health Bureau, Government of the Hong Kong Special Administrative Region (grant no. 20190542)., the Theme-based Research Scheme (Project No. T11-712/19-N) of the Research Grants Council of the Hong Kong Special Administrative Region, China, and the National Institute of Allergy and Infectious Diseases (grant no. R01 AI170116). The original vaccine trial and 5-year follow-up was supported by the Research Fund for the Control of Infectious Diseases of the Health, Welfare and Food Bureau of the Hong Kong SAR Government (grant numbers CHP-CE-03 and 11100882), and the Area of Excellence Scheme of the Hong Kong University Grants Committee (grant number AoE/M-12/06).

