## Supplementary for "A Framework for Measuring Population Immunity Against Influenza Using Individual Antibody Titers"

### 1 Estimators of population immunity

#### 1.1 Geometric mean of titer

The seroprevalence data was stratified into three age groups: 0–17, 18–50, and over 50 years old, and categorized across 10 HAI titer levels (<10, 10, 20, 40, 80, 160, 320, 640, 1280, ≥2560). Serum samples with a titer of <10 were assigned a value of 5 and those with ≥2560 were assigned a value of 2560. Geometric mean HAI titers (GMT) were calculated, weighted by proportion of age groups.

#### 1.2 Proportion of non-naïve individuals

Non-naïve individuals were defined as titer of >10. The proportion of non-naïve individuals was calculated using weights based on the age groups.

#### 1.3 Proportion of population immune

The infection was defined as 4-fold or greater rise in HAI titers. We estimated the associated between pre-epidemic HAI titers and their protection effect against infection through a logistic regression.

$$Y_{4-fold}=\alpha+\beta\cdot{titer}_{pre}$$

We estimated $\beta$ for children and adult age group separately, and obtained the seroprotection level from influenza by the $j$th HAI titer level $z_{jc}$ and $z_{ja}$. We adopted the method by from Cheung et al^1^ for estimation of proportion of population immune and relative reduction in reproductive number.

To estimate the proportion of population in each HAI titer group for each age group, we used Bayesian inference with Dirichlet conjugates for multinomial likelihood$\frac{y_{i}}{x_{i1}!\ldots x_{im}!}\prod_{j=1}^{m} s_{ij}^{x_{ij}}$, where $s_{ij}$ was the proportion of age group $i$ with the $j$th HAI titer, $y_{i}$ was the number of individuals in age group $i$ and $x_{ij}$ was the number of subjects in age group $i$ with the $j$th HAI titer ^2^. We assumed non-informative priors with parameters $\alpha_{j}= 1$ for all HAI titer level j, and hence the joint posterior distributions of $(s_{i1},\ldots, s_{im})$ were Dirichlet distributions with parameters $\alpha_{ij}= x_{ij}+1$ for $j= 1,\ldots,m$. The proportion of the population that was immune could be obtained from $\sum_{i=1}^{n} p_{ij}\sum_{j=1}^{m} s_{ij}z_{j}$, where $z_{j}=z_{jc}$ when the individual is child, or $z_{j}=z_{ja}$.

#### 1.4 Relative reduction in reproductive number

We constructed the next-generation matrix (NGM) $\left\{ Q_{ij} \right\}$, where $Q_{ij}$ represented the average number of cases in age group $i$ generated by a primary infection in age group $j$, through a social contact matrix of Hong Kong^3^. The basic reproduction number R_0_ was defined as the largest eigenvalue of $\left\{ Q_{ij} \right\}$ ^4, 5^. The susceptible proportion of age group $i$ was $1-\sum_{j=1}^{m} s_{ij}z_{j}$, we then constructed another matrix $\left\{ M_{ij} \right\}$, $M_{ij}$= $(1-\sum_{j=1}^{m} s_{ij}z_{j})Q_{ij}$, thus the effective reproduction number R_e_ was the largest eigenvalue of $\left\{ M_{ij} \right\}$. Given that population immunity profile, we calculated the corresponding relative reduction in transmissibility as $1-\frac{R_{e}}{R_{0}}$.

The 95% credible intervals for all parameters estimated were generated using 1000 samples randomly drawn from the joint posterior distribution of $(s_{i1},\ldots, s_{im})$ for each age group.

### 2 Simulation

#### 2.1 Age-HAI titer- structured SEIR model

In our simulation study, we conducted a simulation study using an age-baseline HAI titer-structured SEIR model. We divided the total populations (N = 7150,000) into four age groups 0–17, 18–49, over 50 years old and ten HAI-titer level groups (baseline HAI titer = 0,1, 2, …, 9). Let $N_{ik}$ (k = 1, 2, 3, 4; k = 1, 2, …, 10) represent the population in age group i with HAI titer level k. The number of individuals in each class is given by certain functions of by $S_{ik}\left( t \right)$, $E_{ik}\left( t \right)$,$I_{ik}\left( t \right)$, $R_{ik}\left( t \right)$ respectively. For an age-structured model, we incorporate an age-contact matrix describing the rate of contact between each pair of age-brackets. The contact matrix $C_{ij}$ was estimated based on ^6, 7^.

Moreover, the dynamics of the model are given by the set of ordinary differential equations:

$$\frac{dS_{ik}}{\text{ }dt}=-\beta\cdot{\eta_{ik}S}_{ik}\cdot\sum_{j} C_{ij} \cdot\frac{I_{j}}{N_{j}}$$

$$\frac{dE_{ik}}{\text{ }dt}=\beta\cdot{\eta_{ik}S}_{ik}\cdot\sum_{j} C_{ij} \cdot\frac{I_{j}}{N_{j}}-\sigma\cdot E_{ik}$$

$$\frac{dI_{ik}}{\text{ }dt}=\sigma\cdot E_{ik}-\gamma\cdot I_{ik}$$

$$\frac{dR_{ik}}{\text{ }dt}=\gamma\cdot I_{ik}$$

which depend on the following parameters:

- the total population in j-th age group $N_{j}$;
- the exposed rate $\sigma,$ , given by 1/*L* where *L* is the length of incubation period.
- the recovery rate 𝛾, given by 1/*D* where *D* is the length of the period for which a person is infectious.
- the transmission rate 𝛽, measured as the average number of contacts per person per time; given by R0 * D.

See Table S1 for more details for other parameters.

#### 2.2 HAI titer dynamics model

Denote $T_{i}$ as the infection time for an individual i. Before infection time $T_{i}$, the antibody titers naturally wane over time. We assume the individual decay rate follows a Gamma distribution: $w_{i}\sim Gamma\left( \alpha_{w},1 \right)\text{ }$

$$AT_{i}\left( t \right)=AT_{i}\left( t_{0} \right)*\exp\left( -\frac{w_{i}}{365}*(t-t_{0} \right))$$

The titer will receive a boost after the individual is infected. The titer level at infection time $T_{i}$ is:

$$AT_{i}\left( T_{i} \right)= AT_{i}\left( t_{0} \right)*\exp\left( -\frac{w_{i}}{365}*(T_{i}-t_{0} \right))$$

We assume such boosting follows a Gamma distribution,$\alpha_{i}\sim Gamma\left( \alpha_{b},1 \right)\text{, }$and the boosting takes 14 days.

$$AT_{i}\left( T_{i}+u \right)=AT_{i}\left( T_{i} \right)+\frac{\alpha_{i}}{365}\text{*}\text{ }u , u\leq14$$

We also assume such boosting is temporary and does decay over time. Titer level at $u$ days after infection time is:

$$AT_{i}\left( T_{i}+u \right)=AT_{i}\left( T_{i}+14 \right)*\exp\left( -\frac{w_{i}}{365}*u \right), u>14$$

#### 2.3 Baseline HAI titer distribution

We assume the integer part of the ‘true’ baseline titer follows a multinomial distribution with 10 levels (0 to 9), and the non-integer part follows a uniform distribution. The density of this distribution:

$$f(x)=p_{\left[ x \right]},x\in[0,10)\text{ }$$

### 3 Sensitivity analysis

We set varying protection levels for HAI titer. For setting 1, we set the relative risk reduction is (0, 0.1, 0.2, 0.3, …, 0.9, 1) for 10 HAI titer levels. For setting 2, we set the relative risk reduction is (0, 0.167, 0.333, 0.5, 0.583, 0.667, 0.75, 0.833, 0.917, 1) for 10 HAI titer levels.

**Table S1**. Characteristics of unvaccinated participants with paired sera available for each influenza H1N1 and H3N2 epidemic studied.

| **H1N1** |  | Epidemic 1  2009.07 – 2010.01  (n = 2184) | Epidemic 3  2011.01 – 2011.02  (n = 1661) | Epidemic 5  2013.02 – 2013.04  (n = 1433) | Epidemic 7  2014.01 – 2014.03  (n = 1307) |
| --- | --- | --- | --- | --- | --- |
|  | **Overall** | 2184 | 1661 | 1433 | 1307 |
|  | **Age(years)** |  |  |  |  |
|  | Adult | 1231 | 916 | 782 | 725 |
|  | Child | 953 | 745 | 651 | 582 |
|  | **Sex** |  |  |  |  |
|  | Female | 1170 | 882 | 764 | 688 |
|  | Male | 1014 | 779 | 669 | 619 |
|  | **Received TIV or self-reported vaccination** | 384 | 56 | 163 | 127 |
| **H3N2** |  | Epidemic 2  2010.08 – 2010.10  (n = 567) | Epidemic 4  2012.03 – 2012.06  (n = 1465) | Epidemic 6  2013.07 – 2014.03  (n = 691) | Epidemic 8  2014.06 – 2014.07  (n = 657) |
|  | **Overall** | 567 | 1465 | 691 | 657 |
|  | **Age(years)** |  |  |  |  |
|  | Adult | 316 | 803 | 388 | 365 |
|  | Child | 253 | 664 | 303 | 282 |
|  | **Sex** |  |  |  |  |
|  | Female | 305 | 776 | 370 | 352 |
|  | Male | 264 | 691 | 321 | 305 |
|  | **Received TIV or self-reported vaccination** | 123 | 144 | 75 | 53 |

**Table S2.** Model parameters, descriptions, and values. The total population of these age-group in Hong Kong is 7150,000.

| **Parameter** | **Description** | **Value** | **Notes** | **References** |
| --- | --- | --- | --- | --- |
| $N_{\mathrm{agc}}$ | Number of age groups | 3 |  |  |
| $\beta^{\text{agc }}$ | Disease transmission rate |  |  | Estimated |
| C | $N_{\mathrm{agc}}\times N_{\mathrm{agc}}$ population mixing matrix |  |  | ^3^ |
| $\Phi$ | Vector of the scaling parameters for the contact rates |  |  | Estimated |
| $L$ | Incubation period | 2 |  | ^8^ |
| $D$ | Infectious period | 3 |  | ^8^ |
| $\sigma$ | Exposed rate | 1/2 |  |  |
| $\gamma$ | Infection recovery rate | 1/3 |  |  |
| $R_{e}$ | Effective reproductive number | 1.1,1.2,1.3,1.4 1.5 |  | ^9^, ^10^ |
| $\alpha_{b}$ | Boosting of HAI titer after infection | 78, 104 | $\alpha_{i}\sim Gamma\left( \alpha_{b},1 \right)$  Suppose there would be an 8-fold or 16-fold rise after infection | ^10^ |
| $\alpha_{w}$ | Waning of HAI titer | 0.0027, 0.0018,  0.0012,  0.0007 | $w_{i}\sim Gamma\left( \alpha_{w},1 \right)$  Suppose the waning level of titers 20% to 50% | ^10^ |
| $P_{\text{age }}$ | Population of Hong Kong by age-group | 930,000 (0-17 years) |  | ^7^ ^6^ |
|  |  | 3071,000 (18-49 years) |  |  |
|  |  | 3530,000(50+years) |  |  |

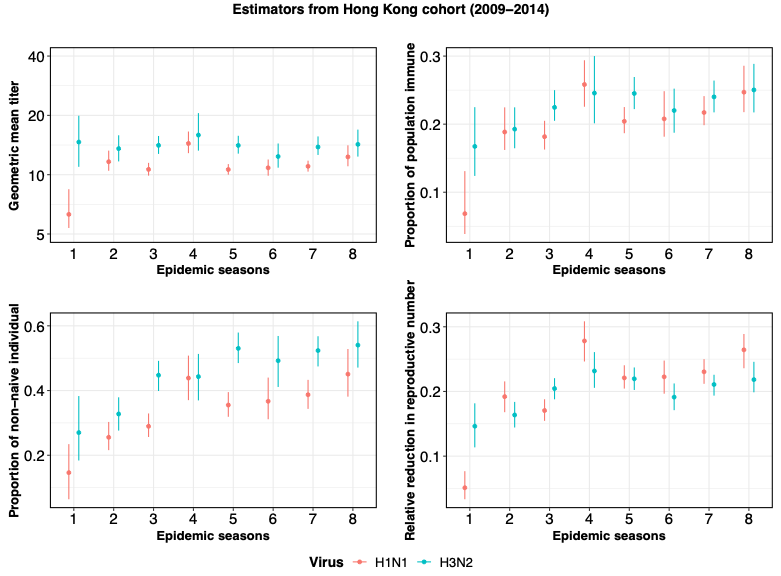

**Figure S1.**

The value of four estimators for influenza H1N1 and H3N2 for the Hong Kong cohort study (2009-2014). Each data point indicates the estimated value. The error bar indicates the 95% confidence interval.

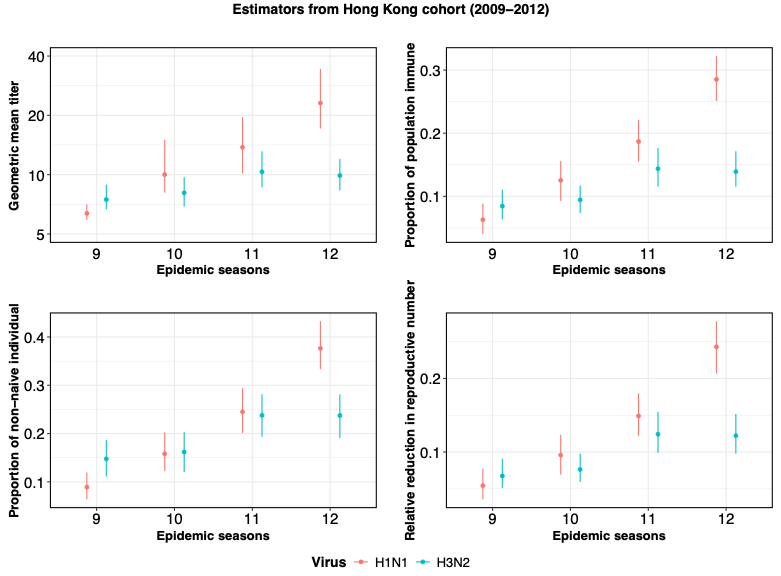

**Figure S2.**

The value of four estimators for influenza H1N1 and H3N2 for the Hong Kong cohort study (2009-2012). Each data point indicates the estimated value. The error bar indicates the 95% confidence interval.

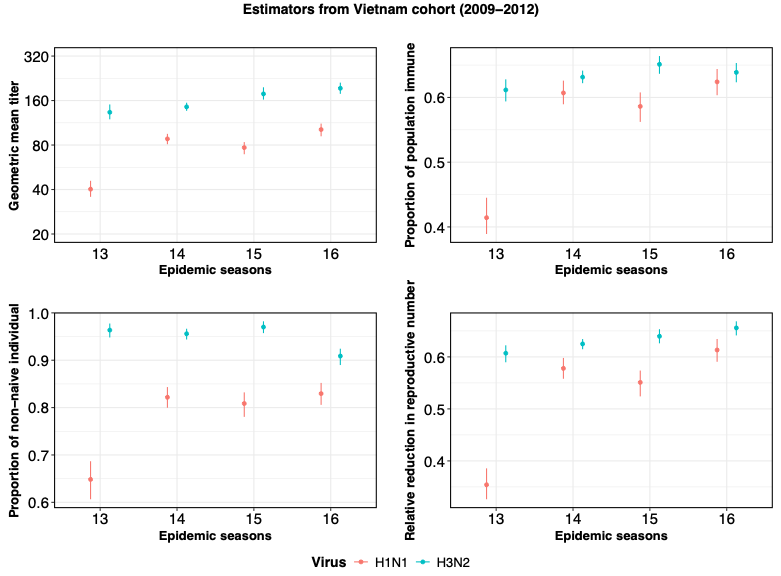

**Figure S3.**

The value of four estimators for influenza H1N1 and H3N2 for Vietnam cohort study (2009-2012). Each data point indicates the estimated value. The error bar indicates the 95% confidence interval.

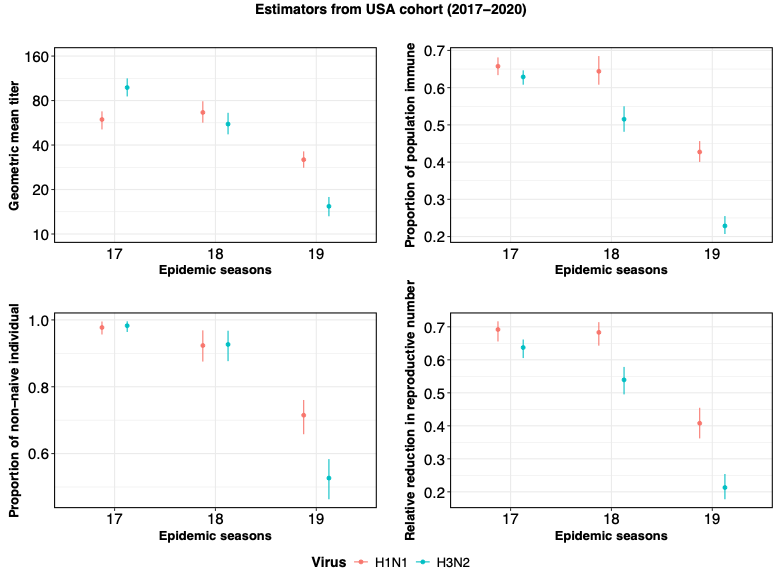

**Figure 4**

The value of four estimators for influenza H1N1 and H3N2 for USA cohort study (2017-2020). Each data point indicates the estimated value. The error bar indicates the 95% confidence interval.

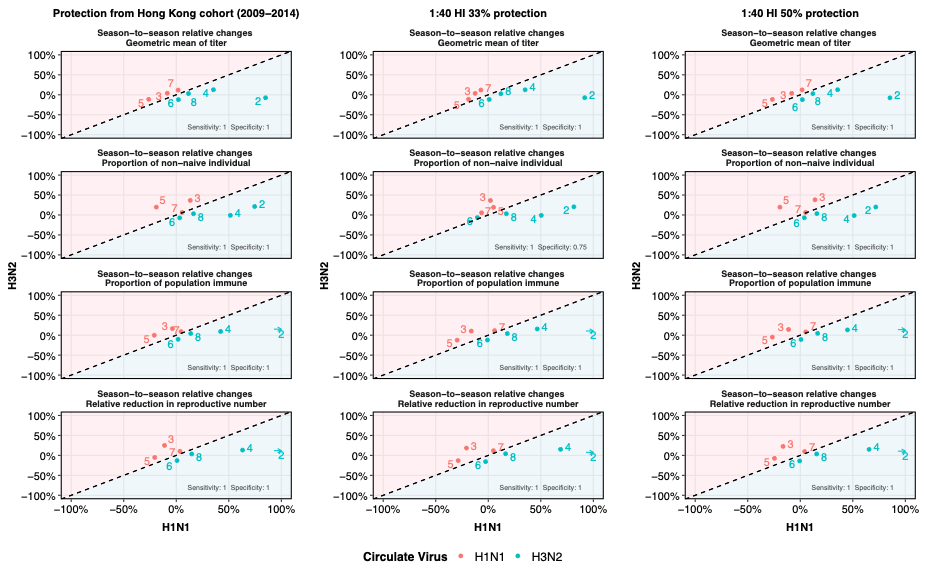

**Figure S5.**

The comparison of relative change in each estimator for H1N1 and H3N2 at different HAI titer- associated protection settings of Hong Kong cohort study (2009-2014).

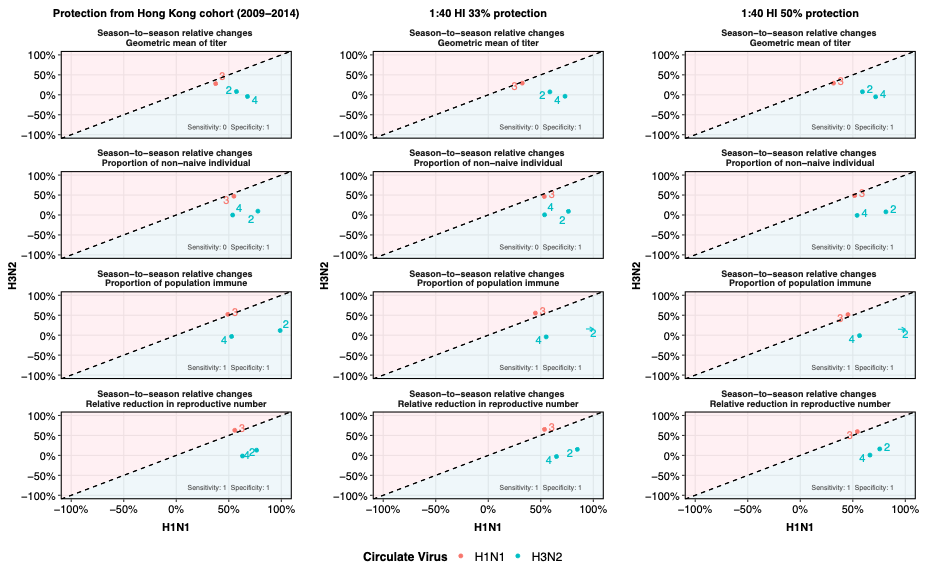

**Figure S6.**

The comparison of relative change in each estimator for H1N1 and H3N2 at different HAI titer- associated protection settings of another Hong Kong cohort study (2009-2012).

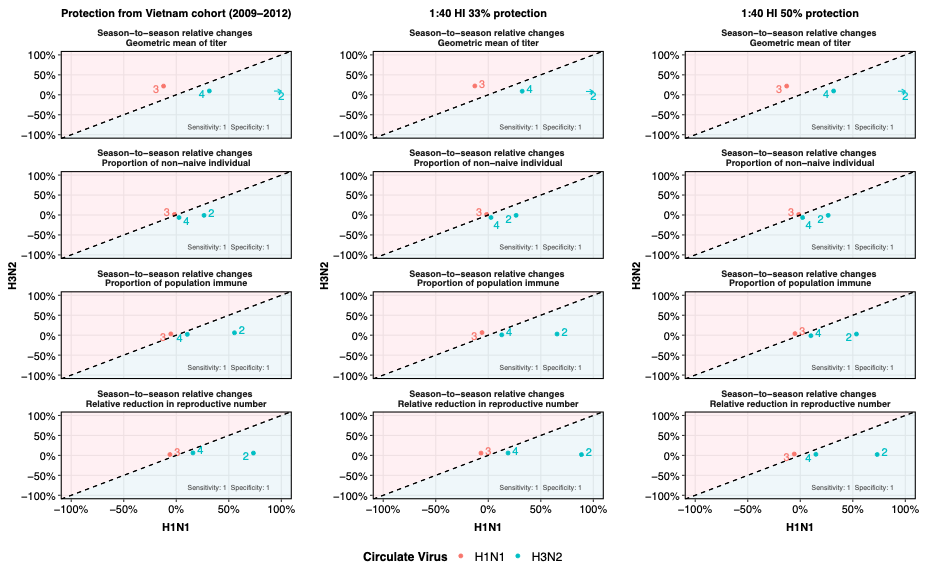

**Figure S7.** The comparison of relative change in each estimator for H1N1 and H3N2 at different HAI titer- associated protection settings of Vietnam study (2009-2012).

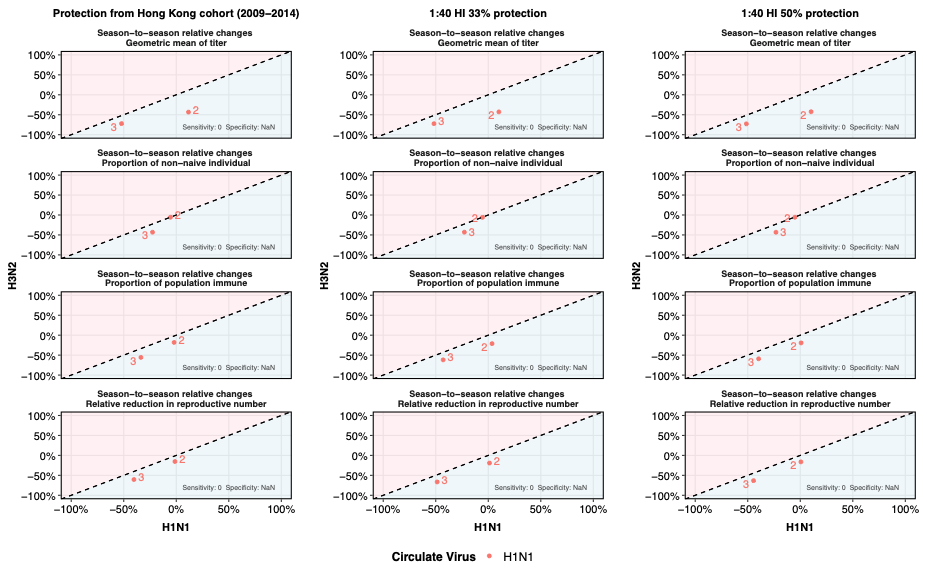

**Figure S8.** The comparison of relative change in each estimator for H1N1 and H3N2 at different HAI titer- associated protection settings of USA study (2017-2020).

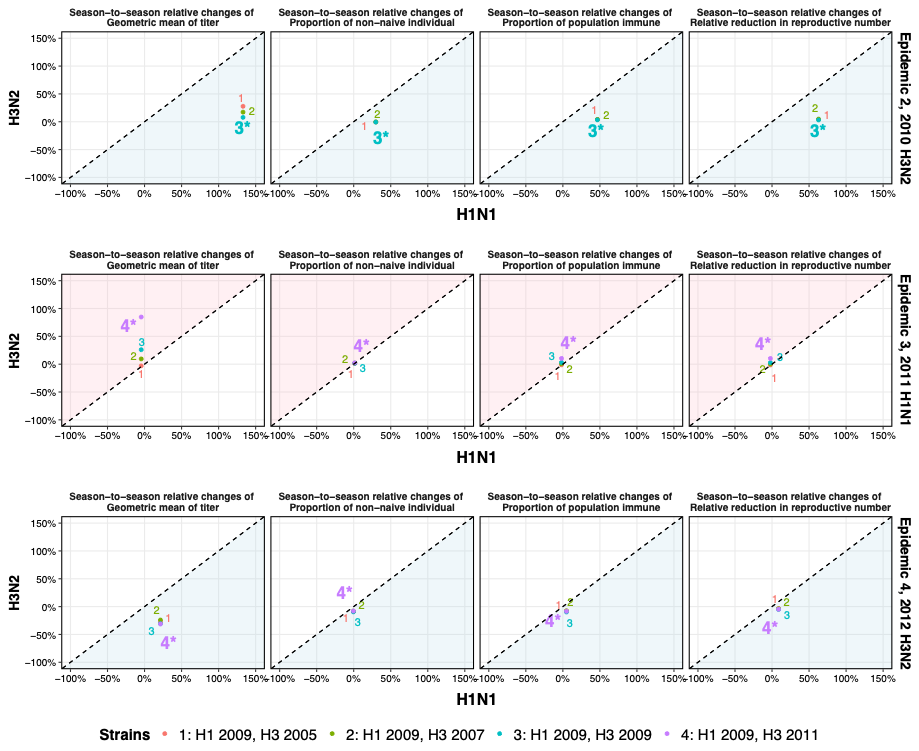

**Figure S9.**

Relative change in each estimator for multiple strains of Vietnam study (2009-2012). The stars indicate the vaccine strains in our primary analysis.

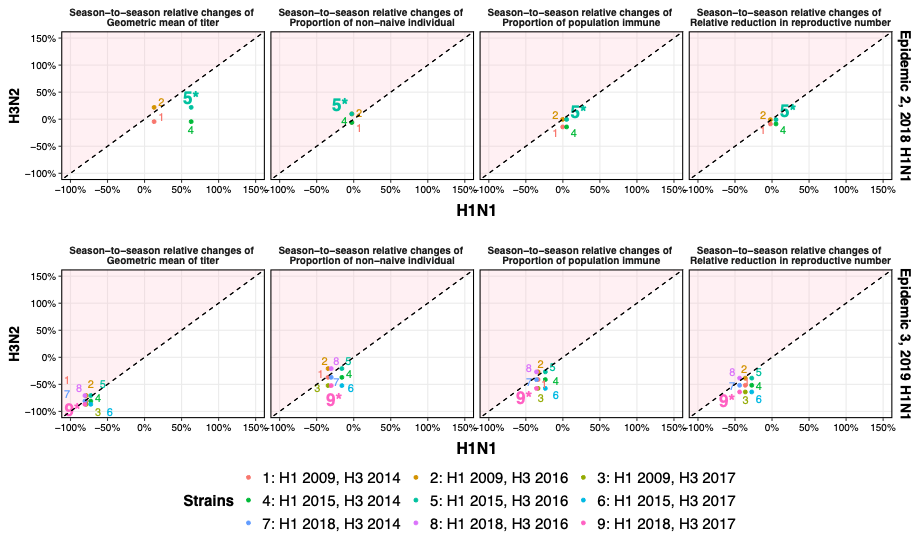

**Figure S10.**

Relative change in each estimator for multiple strains of USA study (2017-2020). The stars indicate the vaccine strains in our primary analysis.

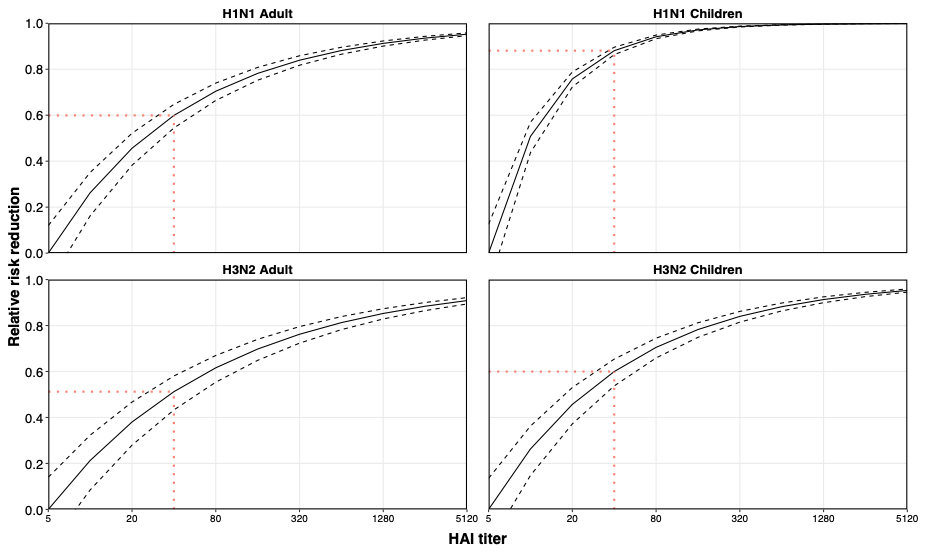

**Figure S11.**

Association between HAI titers and influenza H1N1 and H3N2 virus infection. Solid horizontal and vertical lines show the corresponding 95% credible intervals of the estimates

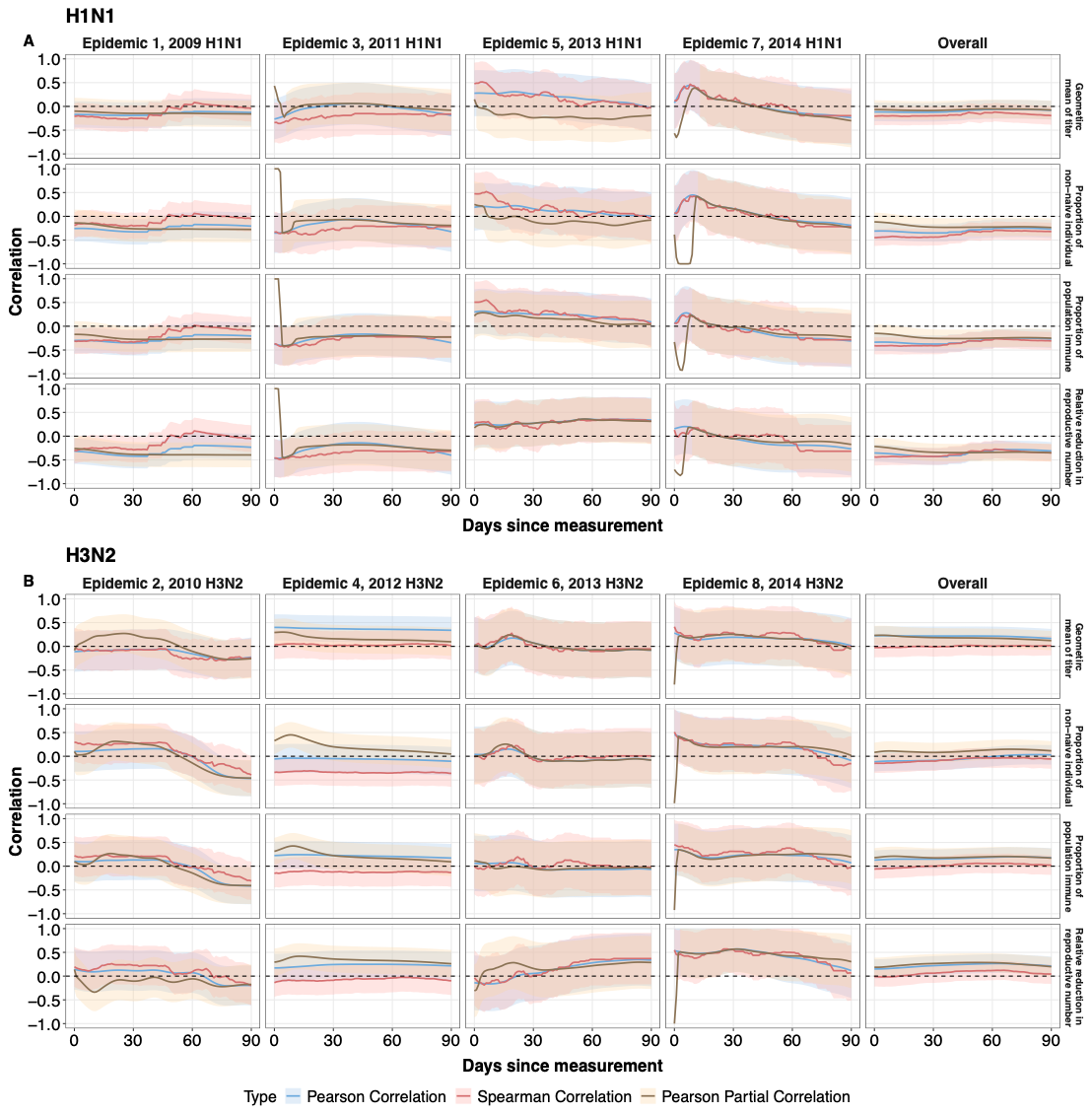

**Figure S12.**

The correlation and partial correlation of current estimators of population immunity on influenza H1N1 and H3N2 activity proxy at few days later for each epidemic season. The blue and red lines show the Pearson correlation and Spearman correlation. The brown line shows the Partial Pearson correlation adjusted for influenza activity at previous 14days. The shaded area represents the 95% confidence interval.

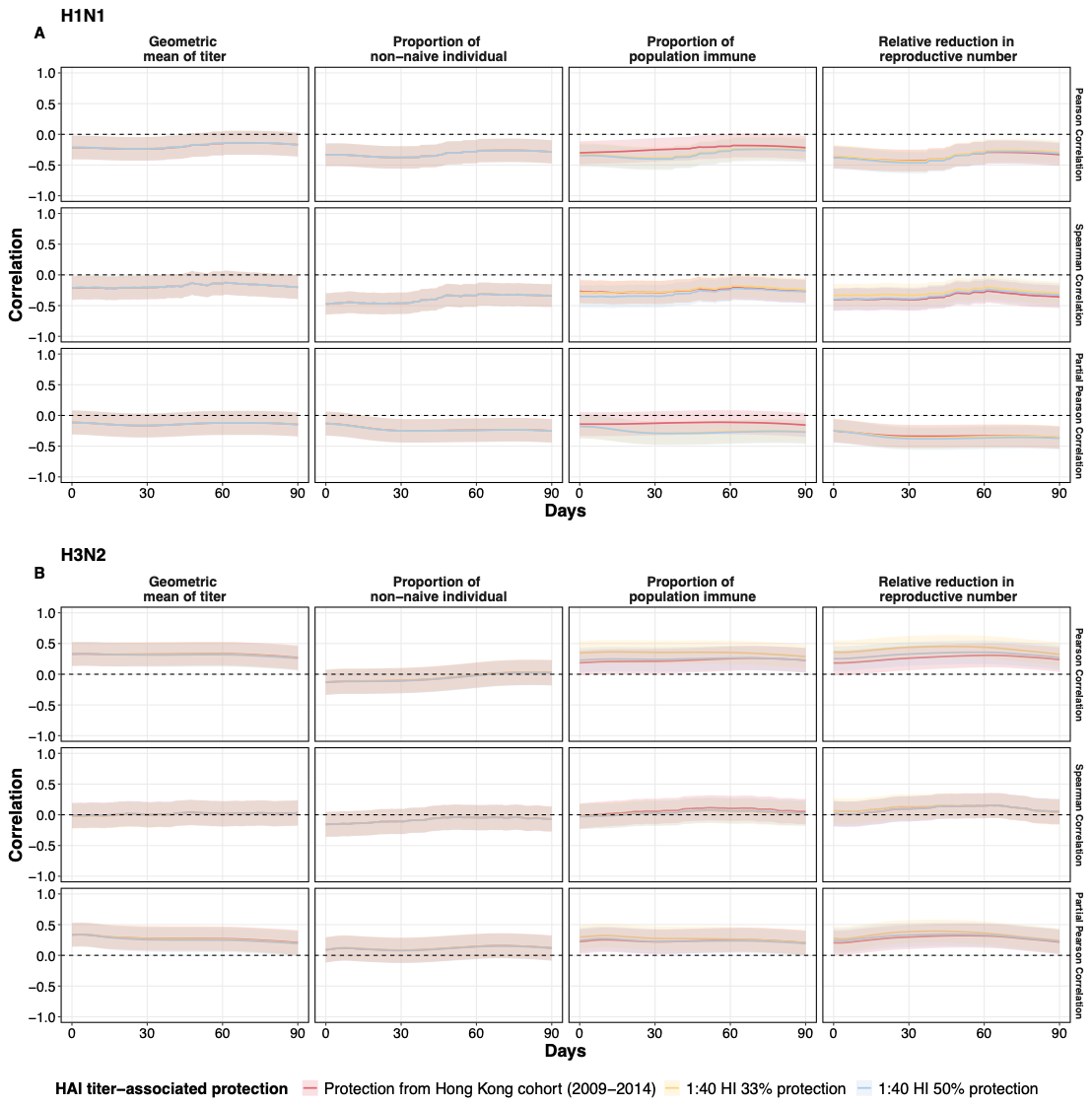

**Figure S13.**

The comparison of correlation and partial correlation between estimators of population immunity and influenza H1N1 and H3N2 activity proxy at different HAI titer- associated protection settings.

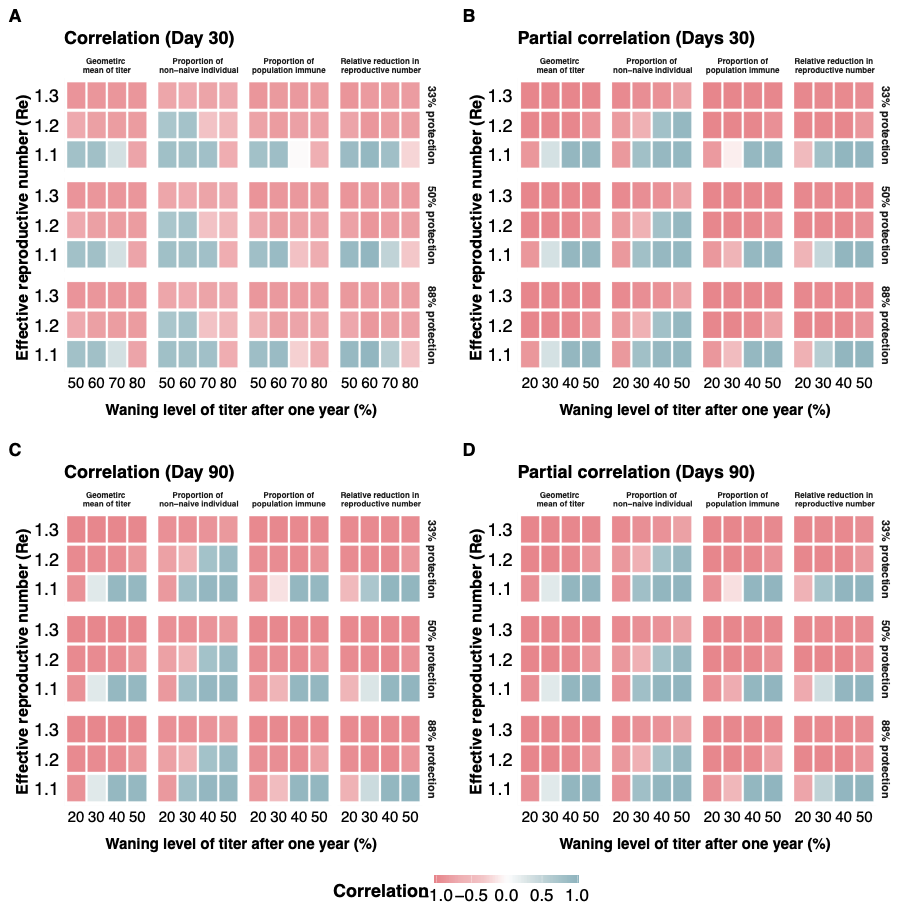

**Figure S14.**

The simulation result for different waning rate, boosting level, effective reproductive number (Re) at different HAI titer- associated protection setting

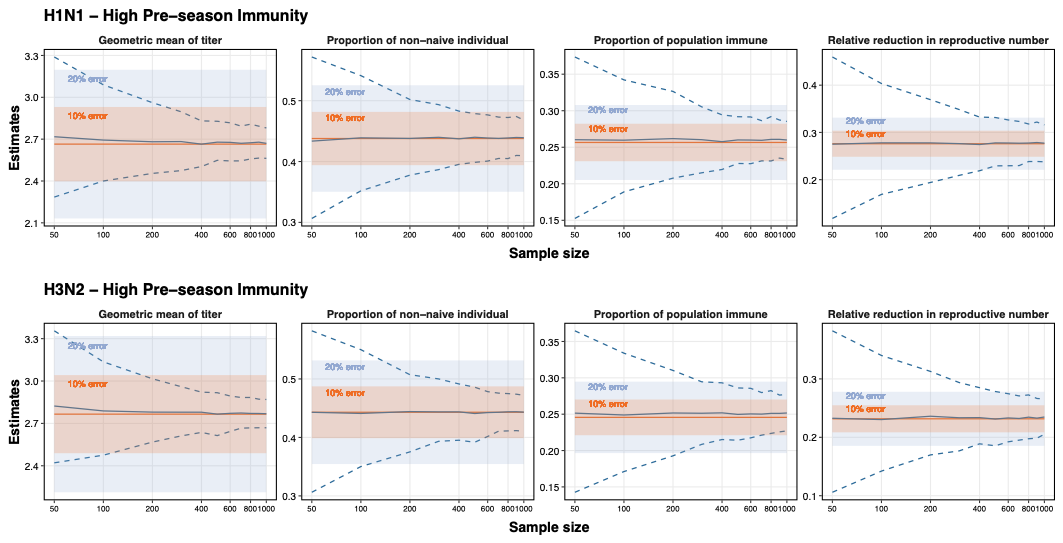

**Figure S15.** Sample size requirements for population immunity estimators with varying precision thresholds for high immunity season. The figure shows the minimum sample sizes required to achieve 10% and 20% error thresholds across four population immunity estimators (geometric mean titer, proportion of non-naive individuals, proportion of population immune, and relative reduction in reproductive number) for H1N1 (top panels) and H3N2 (bottom panels) influenza viruses in average pre-season immunity scenarios.

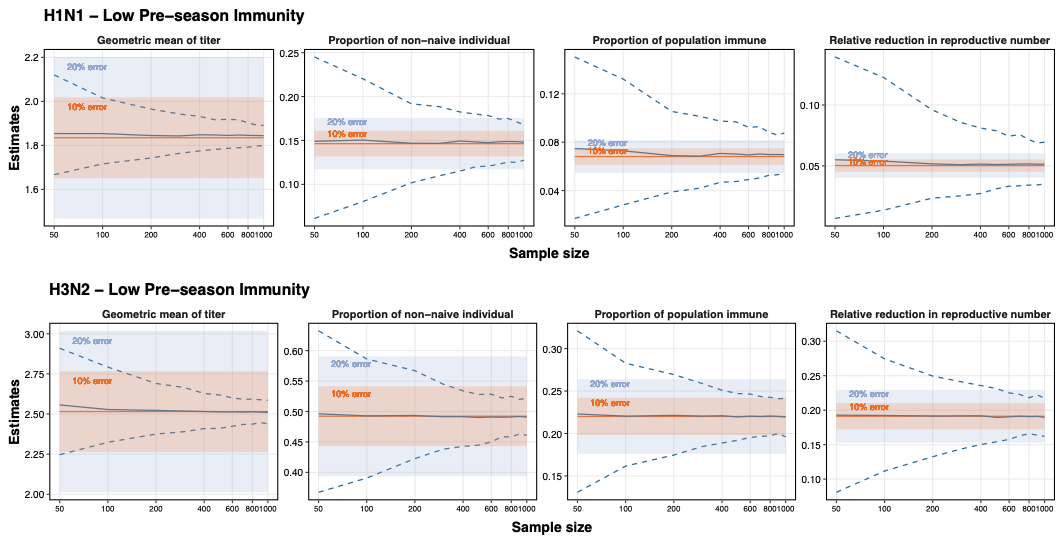
**Figure S16.** Sample size requirements for population immunity estimators with varying precision thresholds for low immunity season. The figure shows the minimum sample sizes required to achieve 10% and 20% error thresholds across four population immunity estimators (geometric mean titer, proportion of non-naive individuals, proportion of population immune, and relative reduction in reproductive number) for H1N1 (top panels) and H3N2 (bottom panels) influenza viruses in average pre-season immunity scenarios.
